# A randomized, double-blind, placebo-controlled clinical study to evaluate the efficacy of the synbiotic medical food, SBD111, for the clinical dietary management of bone loss in menopausal women

**DOI:** 10.1101/2025.05.20.25325893

**Authors:** Eric M. Schott, Mark Charbonneau, Douglas P. Kiel, Susan Bukata, Michael J. Zuscik, Clifford Rosen, Alicia Ballok, Gerardo V. Toledo, Elizabeth Steels, Harry Huntress, Amanda Rao, Thomas G. Travison, Maria J. Soto-Giron, Ian Wolff, D. Davidson Easson, Klaus Engelke, Luis Vitetta

## Abstract

**Summary:** This 12-month study in 286 early postmenopausal women evaluated the efficacy and safety of SBD111, a synbiotic medical food, in reducing bone loss. SBD111 did not significantly reduce bone loss for the full cohort, but did produce evidence of reduced bone loss in women with osteopenia and BMI ≥ 30.

**Purpose:** To determine the efficacy of SBD111, a synbiotic medical food comprising probiotics and prebiotics, in reducing bone loss in women post-menopause, including prespecified subpopulations of women with osteopenia or elevated BMI.

**Methods:** In this prospective, multicenter, double-blind, randomized, placebo-controlled clinical food trial (NCT05009875), 286 healthy, non-osteoporotic women between 1-6 years post-menopause were enrolled and consumed SBD111 (4.75×10^10^ colony forming units) or placebo (maltodextrin) capsules twice daily for 12-months. The primary endpoint was change in areal BMD at the lumbar spine (LS). Secondary endpoints included change in areal BMD at the femoral neck (FN) and total hip (TH), trabecular volumetric BMD at the LS, markers of bone turnover and inflammation, and safety. Changes in gut microbiome composition were exploratory. The hypotheses being tested were formulated before data collection.

**Results:** 286 Women [age 55 ± 3 years (mean ± standard deviation)] were enrolled, with 221 (77%) completing the study. For the primary outcome, SBD111 administration was not associated with significantly less bone loss in the LS after 12-months [0.15% (-0.52%, 0.82%), mean effect size (95% CI) by linear mixed effects regression]. However, SBD111 was associated with reduced BMD loss in the TH for women with BMI ≥ 30 [0.97% (0.015%, 1.925%)] and modestly reduced BMD loss in the FN for women with osteopenia [0.89% (-0.277%, 2.051%)].

**Conclusions:** These findings indicate SBD111 did not significantly reduce BMD loss for the full cohort. However, the trial produced evidence that SBD111 reduced bone loss in women with osteopenia and BMI ≥ 30.

## Introduction

Decreased estrogen production during and after menopause leads to rapid loss of bone mineral density (BMD) throughout the body, rendering women susceptible to osteopenia, osteoporosis, and associated fractures [1]. Osteoporosis and osteopenia affect over 50% of women over age 50, and women with low BMD have a high risk of hip and vertebral fractures, highlighting the need to manage bone loss during and after menopause [2].

Treatments are available to maintain or increase BMD in women, including antiresorptive and osteoanabolic drugs [3], but these therapies are indicated only for patients diagnosed with osteoporosis or at high risk of fracture [4]. Hormone therapy (HT) can be used to manage perimenopausal bone loss, but many women avoid this due to safety concerns [5,6]. Thus, reduction of bone loss is the objective for maintaining skeletal health at the time of menopause, and there is a need to develop new strategies with wide patient acceptance.

Estrogen loss during menopause has effects on bone-resorbing osteoclasts mediated by immune signals [7,8], including tumor necrosis factor alpha (TNF-a), interleukin (IL)-1 Beta (IL-1b), and receptor activator of nuclear factor kB ligand (RANKL) [9]. Blockade of TNF-a or IL-1b has been shown to decrease bone resorption in postmenopausal women [10], and RANKL inhibition with denosumab is commonly used for decreasing bone resorption in patients with osteoporosis [11]. These interventions underscore the importance of systemic inflammation as a driver of bone loss in menopause.

Immunity and inflammation are strongly influenced by the gut microbiota, the diverse community of microbes inhabiting the gastrointestinal tract [12–14]. Probiotic microorganisms can regulate bone remodeling and protect against bone loss through several mechanisms, including the production of anti-inflammatory short chain fatty acids (SCFA) and enhanced gut barrier function [15–17]. Together, these observations highlight an opportunity to identify novel microbial solutions to reduce inflammatory signalling and manage bone loss after menopause.

We previously described SBD111, a synbiotic medical food for the dietary management of postmenopausal bone loss, containing prebiotic dietary fibers and microbial strains isolated from fresh fruits and vegetables [18]. Oral administration of SBD111 significantly reduced bone loss in a mouse model of postmenopausal osteoporosis, and decreased the expression of TNF-a and IL-6 in vertebrae compared to vehicle controls [18]. We also demonstrated that SBD111 is safe and well tolerated in a randomized, placebo-controlled safety and tolerability clinical food trial of 32 healthy adults [19].

Here, we present the results of a clinical food trial designed to test the hypothesis that oral administration of SBD111 will reduce BMD loss in otherwise healthy postmenopausal women over a 12-month period.

## Methods

### Trial design

The study was a multicenter, double-blind, 1:1 randomized, placebo-controlled clinical food trial that enrolled healthy women 1-6 years after menopause. Eligible participants were randomized to receive either an oral medical food, SBD111, or placebo (oral maltodextrin capsules) twice daily for 12-months. Participants’ progression through the trial is presented in **Fig 1** (CONSORT diagram). The visits and measurements schedule is described in **Table SI1** in compliance with SPIRIT guidelines. The study was conducted in compliance with applicable Australian laws and regulations after Human Research Ethics Committee (HREC) and approval from the National Institute of Integrative Medicine (NIIM). Participants were recruited to the study after all necessary approvals had been obtained.

**Fig 1.**
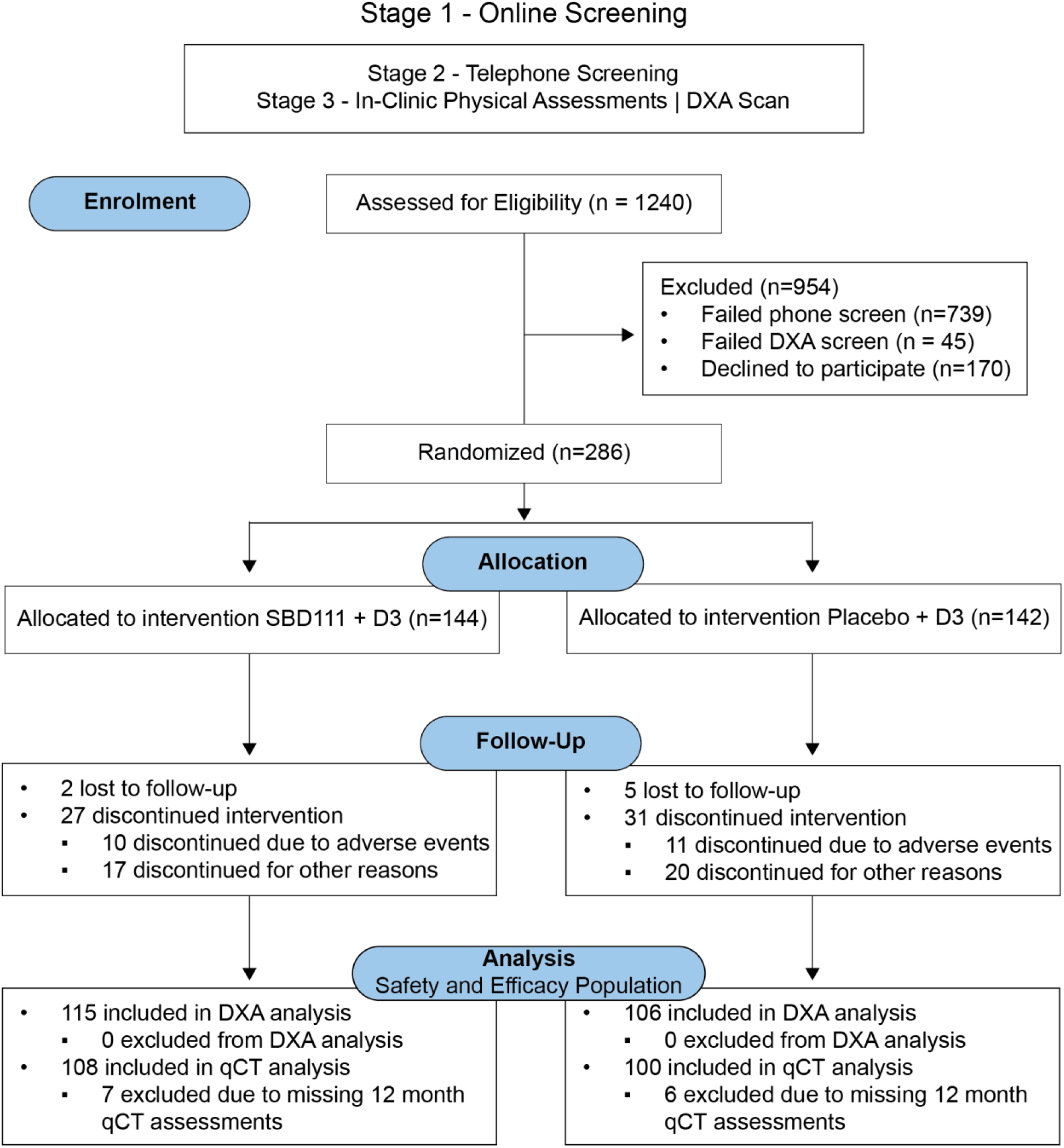
CONSORT diagram depicting the flow of participant screening, enrolment, allocation, follow-up, and analysis during throughout the reported clinical study

### Participant recruitment and allocation

286 women between 1-6 years post-menopause were enrolled across two sites in Brisbane and Sydney, Australia between November 2021 and July 2023. Inclusion criteria included written informed consent, at least 6-months since HT intake, Dual Energy X-Ray Absorptiometry (DXA)-derived T-score >-2.5 at the lumbar spine (L1-L4), femoral neck, and total hip, and BMI between 18.5 and 35 kg/m^2^. Key exclusion criteria included a history of bone disorders, hip replacement, partial hysterectomy, and treatment with osteoporosis drugs (see **Supplemental Information** for all inclusion and exclusion criteria). Eligible participants were randomized to either SBD111 medical food or placebo in a parallel assignment scheme. All participants and study site personnel were blinded to test article allocation. The investigators remained blinded with no access to the randomization code until the end of study, database lock, and statistical analyses were complete.

### Investigational product

The SBD111 synbiotic medical food is composed of Levilactobacillus brevis SBS04254 (3×10^10^ Colony Forming Units (CFU)/day), Lactiplantibacillus plantarum SBS04260 (3×10^10^ CFU/day), Leuconostoc mesenteroides SBS02455 (3×10^10^ CFU/day), Pichia kudriavzevii SBS04263 (5×10^9^ CFU/day), and prebiotics (oligofructose (640 mg/day) and dried blueberry powder (640 mg/day)). The total probiotic concentration was 9.5×10^10^ CFU/day, encased in acid-resistant, delayed-release, size zero, white, opaque capsules (4 capsules/day; **Table SI2**). The placebo was composed of maltodextrin (1,720 mg in 4 capsules/day) in identical capsules. Vitamin D (500IU, NanoCelle D3, Bioglan Medlab) was provided adjunctively to all participants for the duration of the trial.

### Study Outcomes

The primary outcome was the percent change in BMD at the lumbar spine (2-4 evaluable levels L1-L4) after 12-months of administration, attributable to intervention compared with placebo. Other measured outcomes included change in: BMD at the femur and total hip at 6- and 12-months, lumbar spine (L1 & L2) trabecular volumetric BMD (vBMD) and total vBMD at 12-months, serum c-terminal telopeptide (CTX) and procollagen 1 intact N-terminal propeptide (P1NP) at 6- and 12-months, serum inflammatory markers including C-reactive protein (CRP), interleukin-17 (IL-17), tumor necrosis factor alpha (TNFa), interleukin 1 beta (IL-1b), interleukin 4 (IL-4), receptor activator of nuclear factor-κB ligand (RANKL), and interferon gamma (IFNg) at 6- and 12-months, gut microbiome composition and functional potential at 6- and 12-months, and safety and gastrointestinal tolerability (see **Supplemental Information**).

### Prespecified Analytic Subgroups

In addition to the primary and secondary efficacy outcome measures, we prespecified several subgroup analyses: i) duration of time since start the of menopause (1 to 3-years vs. 4 to 6-years), ii) obesity measures at baseline visit (BMI ≥ 30 kg/m^2^ vs. BMI <30 kg/m^2^), iii) presence (T-score £1.0 and >-2.5) vs. absence (T-score >-1.0) of osteopenia in the lumbar spine at baseline.

### Blood samples

Overnight fasted blood samples were collected at enrollment, baseline, 6- and 12-months from all participants. Full blood counts were performed, and serum samples were stored at −80°C before analysis. Serum CTX, P1NP, CRP, IL-17, TNFa, IL-1b, IL-4, RANKL, and IFNg were analyzed by Cardinal Bioresearch Pty. Ltd.

Brisbane, Australia (see **Supplemental Information**).

### Radiology assessment

BMD of the lumbar spine (L1-L4), total hip, and femoral neck of the left femur and body composition were measured by DXA (Lunar Prodigy, GE Medical Systems, or Hologic Horizon, Hologic, Inc) at baseline, 6-, and 12-months. vBMD and lumbar spine architecture were acquired by quantitative computed tomography (QCT) of the lumbar spine (L1 & L2) at baseline and 12-months using whole-body CT scanners (Somatom Definition Edge, Siemens, Inc, Incisive, Philips Healthcare; or Lightspeed VCT, GE Medical Systems) via a custom scanning protocol as previously described [20]. DXA and QCT data were uploaded to a central imaging analysis center (Clario, USA) for image processing, quality control, and analysis by blinded technicians. QCT data were analyzed using Medical Imaging Analysis Framework (MIAF) Spine software (see **Supplemental Information**) [21].

### Statistical Analysis

The study was estimated to have 80% power to detect a difference of 1% in BMD over 12-months with an assumed standard deviation of 2.9% and an estimated evaluable sample of 133 participants per study arm. Anticipating a 10% drop out rate, investigators planned to enroll 300 participants (150 per arm). Efficacy analyses were conducted using the intention-to-treat principle and all available data on randomized participants was included. The primary analysis used mixed-effects linear regression in recognition of the repeated-measures design. The target estimate was the population-level one-year difference in change in BMD attributable to the application of the intervention relative to control, as estimated by a regression contrast at 12-months. Parallel approaches were used for other endpoints. Mixed-effects linear regression models using 6- and 12-month data were supplemented with Students t-tests of the null hypotheses against one-sided alternatives favoring SBD111, motivated by published and unpublished preclinical evidence of consistently reduced BMD loss in animals receiving oral SBD111 across multiple studies [18]. Analyses of serum biomarkers of bone turnover and inflammatory cytokines utilized two-sided Student’s t-tests between groups, as preclinical evidence for these markers was more limited. Adverse and serious adverse events totals were computed for each study arm, as were the proportion of participants experiencing one or more event. See **Supplemental Information** for a comprehensive description of statistical analyses utilized for stool metagenome analysis.

## Results

### Participant recruitment and randomization

286 women age 55±3 years, height 1.7 ± 0.1 meters, and weight 70 ± 12 kg (mean ± standard deviation) were enrolled and randomized to receive SBD111 (n=144) or placebo (n=142) capsules for 12-months, in addition to daily oral Vitamin D_3_ (500IU/day) for the duration of the trial. At randomization, study groups were well balanced across age, height, weight, BMI, blood pressure, and BMD (**Table 1**). 221 participants (77.3%) completed the study and represent the safety and efficacy population for statistical analysis (**Fig 1**).

**Table 1.** Demographics and participant baseline characteristics. Values represent mean (standard deviation)^a^.

| | | All Participants | | Participants with BMI $\geq 30$ at Baseline | | Participants with Osteopenia at Baseline | |
| --- | --- | --- | --- | --- | --- | --- | --- |
| Group |  | SBD111 | Placebo | SBD111 | Placebo | SBD111 | Placebo |
| Enrolled |  | 144 | 142 | 30 | 20 | 52 | 58 |
| Completed |  | 115 | 106 | 26 | 16 | 43 | 40 |
| Withdrew |  | 17 | 20 | 3 | 3 | 6 | 11 |
| Withdrew with AE |  | 10 | 11 | 1 | 1 | 2 | 5 |
| Lost to Follow-up |  | 2 | 5 | 0 | 0 | 1 | 2 |
| Age, years |  | 55 (3.58) | 55 (3.42) | 56 (2.03) | 56 (2.95) | 55 (3.58) | 56 (2.91) |
| Height, meters |  | 1.66 (0.06) | 1.65 (0.07) | 1.66 (0.06) | 1.65 (0.07) | 1.66 (0.06) | 1.66 (0.07) |
| Weight, kilograms |  | 71 (13) | 69 (12) | 89 (7) | 89 (9) | 68 (13) | 68 (12) |
| BMI, kg/m<sup>2</sup> |  | 26 (4.33) | 26 (4.02) | 32 (1.85) | 33 (1.75) | 25 (4.83) | 25 (4.08) |
| Systolic BP, mmHG |  | 124 (15) | 119 (15) | 132 (16) | 127 (14) | 120 (15) | 119 (16) |
| Diastolic BP, mmHG |  | 83 (9.6) | 80 (10) | 87 (7) | 87 (12) | 82 (10) | 79 (10) |
| Baseline BMD, g/cm<sup>2</sup> | Lumbar Spine | 1.08 (0.17) | 1.05 (0.14) | 1.12 (0.96) | 1.07 (0.14) | 0.92 (0.08) | 0.93 (0.08) |
|  | Femoral Neck | 0.84 (0.13) | 0.85 (0.13) | 0.87 (0.12) | 0.87 (0.15) | 0.77 (0.11) | 0.76 (0.09) |
|  | Total Hip | 0.92 (0.11) | 0.92 (0.11) | 0.96 (0.10) | 0.97 (0.12) | 0.86 (0.10) | 0.83 (0.07) |

<sup>a</sup>Osteopenia at baseline defined as a DXA-derived T-score $< -1.0$ in the lumbar spine.

### BMD loss and bone turnover markers for the safety and efficacy population

In the safety and efficacy population (n=221), participants exhibited 91.23 ± 8.19% adherence (mean ± standard deviation), measured by number of capsules remaining at study completion. DXA revealed that BMD decreased at all three skeletal sites examined at 12-months compared to baseline (0.70-1.01% mean BMD decrease per site at 12 months across groups; **Table 2**). For the primary outcome of BMD change at the lumbar spine at 12-months, decline was lower in women receiving SBD111 than those receiving placebo [0.154% (-0.516%, 0.824%) mean difference (95% CI)], but this did not achieve statistical significance (**Fig 2a, Table 3**). Consistently, no significant differences were observed by QCT of the L1-L2 vertebrae for total vBMD, cortical vBMD, subcortical vBMD, or trabecular vBMD between study groups for the safety and efficacy population (**Table SI4**). In addition, no significant differences in change from baseline in serum bone turnover markers CTX and P1NP were observed between groups for this cohort (**Table SI5**).

**Table 2.** Change in BMD measured by DXA for all participants by study group and visit^a^.

|  | **Month 12** |  | **Month 6** |  |
| --- | --- | --- | --- | --- |
|  | **SBD111**<br>**(N = 115)** | **Placebo**<br>**(N = 105)** | **SBD111**<br>**(N = 122)** | **Placebo**<br>**(N = 115)** |
| **Lumbar Spine** |  |  |  |  |
| BMD, % Change | -0.70 (2.76) | -0.76 (2.76) | -0.55 (2.41) | -0.59 (2.40) |
| T-Score, Change | -0.070 (0.25) | -0.069 (0.25) | -0.058 (0.22) | -0.053 (0.21) |
| Z-Score, Change | 0.0057 (0.25) | 0.0052 (0.25) | -0.018 (0.22) | -0.014 (0.21) |
| **Femoral Neck** |  |  |  |  |
| BMD, % Change | -0.74 (2.74) | -0.95 (2.71) | -0.42 (2.60) | -0.79 (2.45) |
| T-Score, Change | -0.052 (0.17) | -0.060 (0.18) | -0.032 (0.17) | -0.049 (0.15) |
| Z-Score, Change | -0.0061 (0.17) | -0.013 (0.18) | -0.0073 (0.16) | -0.024 (0.15) |
| **Total Hip** |  |  |  |  |
| BMD, % Change | -0.89 (2.07) | -1.01 (1.89) | -0.58 (1.54) | -0.049 (0.15) |
| T-Score, Change | -0.052 (0.17) | -0.060 (0.18) | -0.032 (0.17) | -0.024 (0.15) |
| Z-Score, Change | -0.00061 (0.17) | -0.013 (0.18) | -0.0073 (0.16) | -0.60 (1.61) |

<sup>a</sup>Values represent BMD change from baseline [g/cm<sup>2</sup>; mean (standard deviation)].

**Table 3.** Effect of SBD111 administration on BMD by visit for prespecified subgroups as determined by DXA^a^.

|  | N | **Month 12**<br>Mean (95% CI) | LME<br>p-val | t-test<br>p-val |  | N | **Month 6**<br>Mean (95% CI) | LME<br>p-val | t-test<br>p-val |
| --- | --- | --- | --- | --- | --- | --- | --- | --- | --- |
| **Lumbar Spine** |  |  |  |  |  |  |  |  |  |
| All participants | 220 | 0.154 (-0.515, 0.824) | 0.65 | 0.44 |  | 237 | 0.034 (-0.621, 0.690) | 0.92 | 0.46 |
| BMI < 30 | 178 | -0.102 (-0.852, 0.650) | 0.79 | 0.71 |  | 194 | 0.049 (-0.686, 0.784) | 0.90 | 0.44 |
| BMI ≥ 30 | 43 | 1.169 (-0.298, 2.635) | 0.13 | 0.07 |  | 43 | 0.076 (-1.371, 1.523) | 0.92 | 0.46 |
| Osteopenic | 83 | 0.715 (-0.505, 1.939) | 0.26 | 0.25 |  | 90 | 0.339 (-0.859, 1.536) | 0.58 | 0.28 |
| Non-osteopenic | 137 | -0.170 (-0.949, 0.608) | 0.67 | 0.66 |  | 147 | -0.143 (-0.905, 0.619) | 0.71 | 0.65 |
| < 4 years in menopause | 130 | 0.336 (-0.511, 1.184) | 0.44 | 0.28 |  | 142 | 0.152 (-0.674, 0.979) | 0.72 | 0.35 |
| ≥ 4 years in menopause | 88 | -0.021 (-1.089, 1.049) | 0.97 | 0.60 |  | 95 | -0.115 (-1.171, 0.941) | 0.83 | 0.59 |
| **Femoral Neck** |  |  |  |  |  |  |  |  |  |
| All participants | 220 | 0.163 (-0.525, 0.851) | 0.64 | 0.28 |  | 237 | 0.342 (-0.329, 1.013) | 0.32 | 0.13 |
| BMI < 30 | 178 | -0.036 (-0.810, 0.737) | 0.93 | 0.48 |  | 194 | 0.084 (-0.668, 0.836) | 0.83 | 0.38 |
| BMI ≥ 30 | 43 | 1.006 (-0.509, 2.521) | 0.20 | 0.10 |  | 43 | **1.388 (-0.105, 2.881)** | **0.07** | **0.04\*** |
| Osteopenic | 83 | **0.887 (-0.274, 2.044)** | **0.14** | **0.044\*** |  | 90 | 0.295 (-0.837, 1.426) | 0.61 | 0.26 |
| Non-osteopenic | 137 | -0.275 (-1.124, 0.575) | 0.53 | 0.74 |  | 147 | 0.364 (-0.463, 1.191) | 0.39 | 0.18 |
| < 4 years in menopause | 130 | 0.087 (-0.762, 0.935) | 0.84 | 0.38 |  | 142 | 0.275 (-0.548, 1.098) | 0.51 | 0.25 |
| ≥ 4 years in menopause | 88 | 0.272 (-0.857, 1.399) | 0.64 | 0.30 |  | 95 | 0.241 (-0.867, 1.348) | 0.67 | 0.28 |
| **Total Hip** |  |  |  |  |  |  |  |  |  |
| All participants | 220 | 0.058 (-0.410, 0.526) | 0.81 | 0.34 |  | 237 | 0.019 (-0.437, 0.474) | 0.94 | 0.46 |
| BMI < 30 | 178 | -0.164 (-0.696, 0.368) | 0.55 | 0.62 |  | 194 | -0.127 (-0.643, 0.389) | 0.63 | 0.70 |
| BMI ≥ 30 | 43 | **0.970 (0.015, 1.925)** | **0.05** | **0.041\*** |  | 43 | 0.557 (-0.385, 1.499) | 0.25 | 0.11 |
| Osteopenic | 83 | 0.280 (-0.482, 1.040) | 0.47 | 0.22 |  | 90 | -0.190 (-0.931, 0.550) | 0.62 | 0.71 |
| Non-osteopenic | 137 | -0.045 (-0.627, 0.537) | 0.88 | 0.49 |  | 147 | 0.156 (-0.411, 0.723) | 0.59 | 0.28 |
| < 4 years in menopause | 130 | 0.294 (-0.320, 0.907) | 0.35 | 0.18 |  | 142 | -0.097 (-0.693, 0.499) | 0.75 | 0.64 |
| ≥ 4 years in menopause | 88 | -0.339 (-1.044, 0.363) | 0.35 | 0.74 |  | 95 | 0.066 (-0.627, 0.759) | 0.85 | 0.41 |

<sup>a</sup>Month 12 and Month 6 values represent effect size estimates and 95% confidence interval in percent change of BMD as determined by linear mixed effects models. Positive values correspond to decreased bone loss in SBD111 recipients. LME p-values represent linear mixed effects models-based treatment effect at 6- or 12-months. T-test p-values represent supporting analysis by one-sided Student's t-test, pooled variance.

**Fig 2.**
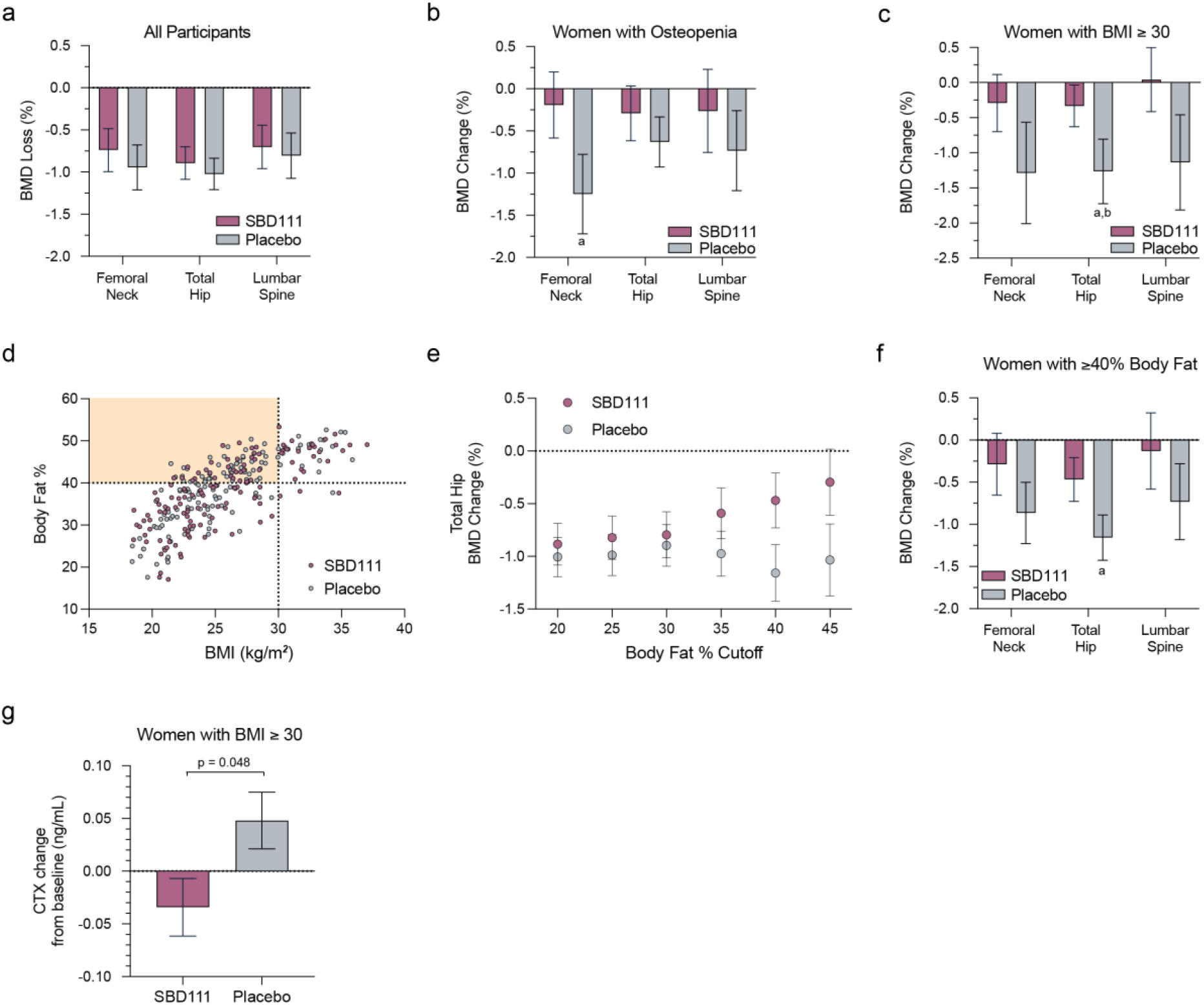
BMD change from baseline for participants with osteopenia, elevated BMI, or elevated adiposity. (a-c) BMD change from baseline to 12 months represented as percent of baseline BMD at the femoral neck, total hip, and lumbar spine for (a) all participants, (b) participants with osteopenia and (c) participants with BMI ≥ 30 that received either twice daily SBD111 or maltodextrin placebo. (d) Scatter plot showing the relationship between BMI (kg/m^2^) and Body Fat % at baseline for participants enrolled in the clinical study. Points represent individual participants, shaded by intervention group (SBD111 or Placebo). (e) BMD change from baseline to 12 months represented as percent of baseline BMD at the total hip for participants that received either twice daily SBD111 or maltodextrin placebo stratified by increasing levels of body fat percentage at baseline. (f) BMD change from baseline to 12 months represented as percent of baseline BMD at the femoral neck, total hip, and lumbar spine for participants with >40% body fat at baseline that received either twice daily SBD111 or maltodextrin placebo. (g) Change in serum CTX concentration from baseline to 12 months for participants with BMI ≥ 30 that received either twice daily SBD111 or maltodextrin placebo. P-value represents a two-sided Student’s t-test. For (a-c) and (e-g), values represent mean ± standard error. ^a^ p < 0.05 one-sided student’s t-test, ^b^ p = 0.05, linear mixed effects model treatment effect at 12 months

### BMD loss and markers of bone turnover and inflammation in prespecified subgroups

Although no significant effects were observed in the full safety and efficacy population, there was evidence of less bone loss in the SBD111 group by DXA in two populations: women with osteopenia at baseline and women with BMI ≥ 30 (**Fig 2b-c, Table 3**). Among osteopenic women (n=83), participants consuming SBD111 lost significantly less BMD in the femoral neck at 12-months than those consuming placebo [0.887% (-0.277%, 2.051%) mean difference (95% CI) by linear mixed effects regression; p=0.044, one-sided student’s t-test] (**Fig 2b, Table 3**). Similar nonsignificant trends were observed at the lumbar spine and total hip (**Table 3**).

Women with BMI ≥ 30 at baseline (n=42) who were assigned to the SBD111 group also exhibited reduced BMD loss in the total hip at 12-months [0.970% (0.015%, 1.925%) mean difference (95% CI) by linear mixed effects regression; p = 0.041, one-sided student’s t-test] (**Fig 2c, Table 3**) compared to placebo. Similar nonsignificant trends were observed at the lumbar spine and femoral neck at 12-months (**Table 3**).

Importantly, participant characteristics were comparable for women assigned to the SBD111 and placebo arms for these subgroup analyses (**Table 1**).

No significant differences in change of CTX or P1NP from baseline between SBD111 and placebo groups were observed for participants with osteopenia at baseline (**Table SI5**). However, SBD111 administration was associated with a significant decrease in serum CTX compared to placebo controls at 12-months in participants with BMI ≥ 30 (p = 0.049, two-sided Student’s t-test; **Fig 2g, Table SI5**).

Obesity is associated with systemic inflammation, driven in part by endocrine and pro-inflammatory mediator production by adipose tissue, and we previously reported that administration of SBD111 reduced inflammatory biomarkers in mice [18,22]. We aimed to determine whether SBD111 administration was associated with changes in serum inflammatory markers in women with BMI ≥ 30. Trends of decreased IL-17A, TNFa, and IL-1b, were observed for participants receiving SBD111 compared to placebo, but these differences did not achieve statistical significance (p > 0.05, two-tailed student’s t-tests, **Fig SI1**).

No significant differences in BMD loss were observed in subgroups of women with BMI < 30, women without osteopenia at baseline, women less than four years after menopause, or those at least four years after menopause (**Table 3**). In addition, no significant differences were observed by QCT of the L1-L2 vertebrae for any metric between study groups for any prespecified study subgroup (**Table SI4**).

### BMD loss in women with elevated body fat

The usefulness of BMI as a metric is debated because it fails to distinguish between lean mass and fat mass. Indeed, 87 women who completed this study (30.4%) exhibited ≥ 40% body fat despite having a BMI ≤ 30 (**Fig 2d**). Further, exploratory analysis revealed that administration of SBD111 was associated with reduced bone loss at the total hip compared to placebo for women with ≥ 40% body fat (n=130), regardless of BMI (**Fig 2e**). This effect was statistically significant at the total hip at 12-months (p=0.035, one-sided student’s t-test), and similar nonsignificant trends were observed at the lumbar spine and femoral neck (**Fig 2f**). However, no significant differences were observed in serum CTX or P1NP for women with ≥ 40% body fat (**Table SI5**).

### Effects of SBD111 administration on the gut microbiota

To characterize the effects SBD111 on the structure and function of the gut microbiota, stool samples underwent metagenomic DNA sequencing. This analysis revealed that SBD111 administration was associated with significant enrichment of SBD111 strains and modestly increased richness of functional pathways in the gut metagenome at 12-months compared to placebo (**Figs SI2-SI3**; see **Supplemental Information**).

Previous clinical studies have reported decreased gut microbial diversity in obese participants [23]. Accordingly, we observed that women with BMI ≥ 30 exhibited significantly reduced species richness (p=0.02, Welch’s t-test) and a trend of reduced Shannon diversity indices (p=0.08) in their stool metagenomes at baseline compared to those with BMI < 30 (**Fig SI4a-b**). Moreover, women with BMI ≥ 30 displayed significantly reduced richness of Metacyc functional pathways, though no significant difference was observed for Shannon diversity of functional pathways (p=0.02 and p=0.83, respectively, Welch’s t-test; **Fig SI4c-d**).

29 bacterial species exhibited significantly lower abundance in women with BMI ≥ 30, including several representatives of genera shown to produce the anti-inflammatory SCFA, butyrate, including Clostridium, Butyrivibrio, and Anaerostipes (**Table SI6**). Similarly, women with ≥ 40% body fat exhibited lower species richness and Shannon indices (p=0.0003 and p=0.0017, Welch’s t-test, respectively; **Fig SI4e-f**), Metacyc functional pathway richness (p=0.016; **Fig SI4g**) and abundance of Clostridial species associated with butyrate production (**Table SI7**) compared to women with body fat < 40%.

### Gastrointestinal tolerability of SBD111

To evaluate gastrointestinal tolerability, participants completed 12-item gastrointestinal tolerability questionnaires (GITQ) on weeks 1-4, as well as at months 3, 6, 9, and 12. 30.3% of participants reported experiencing gastrointestinal (GI) symptoms in at least one survey, indicating that most participants experienced no GI symptoms. No significant differences were observed in the number of participants reporting symptoms or in the total number of GI symptoms reported per participants for mild or moderate symptoms between study groups (p > 0.05, Pearson’s Chi^2^ test; **Table SI8**). However, a significantly lower proportion of participants in the SBD111 group reported experiencing severe GI symptoms (p = 0.02, Pearson’s Chi^2^ test; **Table SI8**). Similarly, participants allocated to the SBD111 group reported a significantly lower number of severe GI symptoms compared to participants randomized to placebo (p = 0.02, Pearson’s Chi^2^ test; **Table SI8**).

### Adverse Events

126 total adverse events (AEs) were reported, with 87 (69%) determined as possibly related to the interventions (SBD111 or placebo) (**Table SI3**). No significant differences were observed in number of AEs reported per participant between study groups or in the frequency of AEs possibly related to study intervention between study groups (p >0.05, Pearson’s Chi^2^ test, respectively; **Table SI3**).

## Discussion

Bone loss remains a significant challenge during and after menopause, and there are few safe and effective solutions to maintain bone mass during this period. In the present study, we observed no significant differences in our primary outcome of lumbar spine BMD between active and placebo groups after 12-months for the full safety and efficacy population. However, in our pre-specified subgroup analyses of women with osteopenia and of those with elevated BMI, SBD111 protected women against bone loss to a significantly greater extent than placebo. Exploratory analysis of women with elevated body fat composition similarly revealed evidence of decreased bone loss in women randomized to the SBD111 as compared to placebo.

Managing bone loss in women with osteopenia is a key unmet need, as their fracture risk is elevated compared to individuals with healthy BMD^3^, and management options are limited. The present study suggests that women with osteopenia could manage bone loss by consuming a medical food synbiotic earlier than they would be eligible for pharmaceutical interventions, potentially reducing their lifetime fracture risk. However, fully evaluating the long-term benefits of the SBD111 medical food for women with osteopenia will require additional clinical investigation over several years of administration in large study cohorts.

The efficacy of SBD111 in obese women is supported by the multiple skeletal outcomes that we examined in this trial. First, administration of SBD111 in women with BMI ≥ 30 was consistently associated with reduced mean BMD loss at three sites: lumbar spine, total hip, and femoral neck. These BMD differences achieved marginal statistical significance only in the total hip, possibly due to the smaller number of participants in this group. Secondly, the observed attenuation of bone loss in women with BMI ≥ 30 kg/m^2^ receiving SBD111 was accompanied by significantly reduced serum CTX, a marker of osteoclast activity and bone loss, as well as trends of reduced serum concentrations of IL-17A, TNFa, and IL-1b. Lastly, metagenome analysis revealed that women with BMI ≥ 30 harbored significantly reduced gut microbial diversity at baseline than those with BMI < 30, characterized by reduced abundance or absence of species known to produce anti-inflammatory SCFA and suggesting a more pro-inflammatory gut environment. This triangulation of evidence supports a true effect of SBD111 on skeletal health in the early menopausal period among women with obesity.

Approximately 43.3% of women over age 40 years in the United States have obesity and could benefit from new solutions for managing postmenopausal bone loss [24]. However, the relationship between elevated BMI and bone health is controversial [25]. Many studies have reported an association between obesity and increased BMD, and this relationship is often attributed to increased mechanical loading on the skeleton [22,26]. Conversely, a study of over 6,000 individuals reported a negative correlation between body fat percentage and bone mass, and a recent meta-analysis found that abdominal obesity is not protective against fractures [27,28]. These findings highlight the complexity of interactions between excess body weight and/or fat and bone health.

Obesity is associated with a state of chronic, systemic inflammation, thought to be driven by endocrine and pro-inflammatory mediators produced by excess adipose tissue [22]. This systemic inflammation is correlated with increased bone loss and fracture [8,29]. Visceral adipose tissue, in particular, produces high levels of TNFa and IL-6 and promotes lower trabecular bone volume, slower bone formation, and higher cortical bone porosity [30,31]. Further, obesity is associated with reduced gut microbial diversity that can impair gut-barrier integrity and potentiate local and systemic inflammation [32]. Taken together, these observations suggest a link between obesity-associated inflammation, the anti-inflammatory properties of SBD111, and evidence of reduced bone loss for postmenopausal women with obesity or those with > 40% body fat receiving the medical food.

This study provides evidence that SBD111 is safe, well tolerated, and reduces bone loss in women with osteopenia and those with BMI ≥ 30, groups that comprised 50.3% of enrolled participants. Further investigation is warranted to fully characterize the effects of SBD111 in these contexts, to determine the mechanisms through which SBD111 can preserve bone mass, and to evaluate long-term efficacy. Importantly, these results provide evidence that synbiotic medical foods, such as SBD111, provide an additional opportunity for the dietary management of bone loss in early menopause.

### Strengths and Limitations

This study has several key strengths. First, it utilized a double-blind, randomized, and placebo controlled design to minimize bias in the study results. Secondly, the study included participants in demographics with elevated risk for bone loss or fracture, including women with osteopenia and those with elevated BMI, and the randomization procedure used ensured comparability between intervention and placebo groups across these prespecified analytic subgroups. Lastly, this study enabled examination of the effects of the synbiotic medical food intervention across several dimensions, including BMD measurements, serum biomarkers of bone turnover and inflammation, and assessment of gut microbiome structure and function. However, this study also has limitations. Most notably, while the safety and efficacy cohort was well powered to detect changes in BMD, the number of participants included in prespecified analytic subgroups is small relative to the effect sizes expected in BMD change. This necessitates confirmatory clinical studies in these populations to validate the results reported here. In addition, this study was conducted in a single country, and demonstrating that these findings generalize to other populations of women would require further investigation.

## Institutional Review Board Statement

The study was conducted in accordance with the Declaration of Helsinki, and approved by the National Institute of Integrative Medicine Human Research Ethics Committee (EC00436) in Brisbane, QLD, Australia (Reference Number: 0089E_2021, August 26, 2021).

## Informed Consent Statement

Informed consent was obtained from all subjects involved in the study.

## Data Availability Statement

Any additional information required to reanalyze the data reported in this paper is available upon request by

