## Supplemental File #1 for "A randomized, double-blind, placebo-controlled clinical study to evaluate the efficacy of the synbiotic medical food, SBD111, for the clinical dietary management of bone loss in menopausal women"

**Supplemental Methods**

*Clinical Trial sites and subject recruitment*

Patient enrollment for the study commenced in March 2022 in Brisbane, QLD Australia. RDC Clinical (Newstead) was the primary study site that managed the participant trial-phase including screening, recruitment, enrollment, collection and storage of blood specimens, administration of the participant reported outcomes, adverse effects, use of rescue medication, progress and trial close out. A single radiology site was selected for DXA and QCT radiological assessment (Brisbane). A central imaging analysis center, Clario (USA), was selected to provide training, quality control, and data analysis of all radiology data. Finally, Microba (Brisbane) was selected to provide microbiome sample kits and central microbiome analysis. In February 2023, additional radiology sites in Brisbane and Sydney NSW were added to facilitate improved participant enrollment. An additional pathology clinic was selected for collection of blood samples in Sydney. The trial was conducted using a purpose built E-Platform for participant management through the trial. Participants were recruited and randomized (n=286) in both Brisbane and Sydney through social media channels, radio advertising and medical clinics. Participant screening and enrollment (**Fig 1**) was overseen by the principal and associate investigators and performed by trained study research associates holding certifications in Good Clinical Practice. A screening log documented all eligible and ineligible persons screened along with reasons for any exclusions. Those who were eligible and recruited to the trial provided written informed consent prior to study enrollment.

Inclusion Criteria

To be eligible for inclusion in the trial, a participant was required to fulfill all of the following criteria:

- i) Provide written informed consent;
- ii) State availability throughout entire study period (12-months) and willingness to fulfill all details of the protocol, including undergo DXA scans (x3), undergo QCT scans (x2), provide stool samples for analysis of gut microbiome (x3), and have blood tests (x3);
- iii) Between 1 – 6 years post-menopause, i.e., at least 1 year but up to a maximum of 6-years since the last menstruation or since having a total hysterectomy;
- iv) At least 6-months since the last intake of hormone therapy (HT);
- v) DXA-derived bone mineral density (BMD) T-score of greater than -2.5 at the lumbar spine (L1-L4), femoral neck, and total hip;
- vi) Body mass index between 18.5 and 35 kg/m$^2$;
- vii) Normal levels of serum calcium (<11mg/dL);
- viii) Normal cardiovascular parameters (systolic blood pressure ≤ 155 mm Hg, diastolic blood pressure ≤ 95 mm Hg), either healthy or medication controlled.

Exclusion Criteria

The presence of any of the following excluded prospective participants from enrolling in the study:

- i) History of other bone disorders (e.g., Paget's disease, or osteomalacia, osteogenesis imperfecta, osteopetrosis, etc.);
- ii) History of cancer treatment with radiation therapy, anti-estrogen therapy, hormonal therapy, or aromatase inhibitors;
- iii) History of bone or colorectal cancer;
- iv) History of autoimmune disorders i.e., rheumatoid arthritis, Hashimoto's disease, Graves' disease, etc, type 2 diabetes mellitus, specific gastrointestinal disorders i.e., ulcerative colitis, Crohn's disease, inflammatory bowel disease, irritable bowel syndrome, kidney disease or dysfunction or any other medical condition that could interfere with the conduct of the study;
- v) History of chronic antibiotic use;
- vi) History of bariatric surgery;
- vii) History of partial colectomy;
- viii) Women with spine abnormalities that would prohibit assessment of BMD;
- ix) Women who have undergone hip joint replacement;
- x) Women who have undergone a partial hysterectomy;
- xi) Women with untreated hyperparathyroidism;
- xii) Women previously treated with calcitonin, estrogens, estrogen derivatives, selective estrogen receptor modulators (SERMs), tibolone, progestins, anabolic steroids, or daily glucocorticoids in the past 6 months;
- xiii) Women treated with bisphosphonates or strontium in the past 5 years;
- xiv) Women previously treated with parathyroid hormone, parathyroid hormone analogs, gallium nitrate, romosozumab or denosumab;
- xv) Per-oral use of corticosteroids;
- xvi) Smoking or use of nicotine products within the past 6-months;
- xvii) Any disease, that by the investigator's judgement, could interfere with the intestinal barrier function;
- xviii) Participation in other bone, diet, autoimmune, or gastrointestinal related clinical trials in the last 6 months;
- xix) Desire and/or plans on changing current diet and/or exercise regime during the participation of this trial;
- xx) Pregnancy or lactation;

xxi) Consumption of dietary supplements (probiotics, prebiotics) in the month prior to or during study (if participant is willing to stop consumption for 1-month, they could be enrolled after a 1-month washout period);

xxii) Prescribed antibiotics in the past 2 months (if participant was placed on an antibiotic after enrollment in the study, the participant was subject to a per protocol analysis).

Changes to Eligibility Criteria During Study

In June 2022, as recruitment was slower than anticipated, the following changes were made to the protocol in an effort to increase recruitment.

1. Instead of exluding women with a history of all cancers except for skin cancer, we instead excluded women with a history of bone or colon cancer. Other history of cancer were eligible if they had been in remission for at least 5-years.
2. Excluded women with a history of cancer and were treated with radiation therapy, anti-estrogen therapy, hormonal therapy, or aromatase inhibitors.
3. Included women with a BMI of 35 or below instead of 32.5 or below.
4. Instead of excluding women who have undergone any joint replacement surgery, we instead excluded only women with a history of a hip joint replacemet surgery.

*Subject Allocation*

Eligible participants were randomized to either the SBD111 medical food group or placebo group without stratification using computer-generated random numbers (Sealed Envelope Software) in a parallel assignment scheme. All participants and study site personnel were blinded to test article allocation, hence with quadruple masking (i.e., participant, care provider, investigators, outcomes assessors) and with documentation blinding. Participant unblinding was only requested in a medical emergency, where knowledge of the study arm was essential for any treatment of the participant. Any reason for unblinding was documented, and the study test article was not revealed to any member of the study team.

*Instructions to participants*

Study participants were instructed to: (i) Consume two capsules twice per day (with breakfast and dinner) with water, (ii) store the study product in the refrigerator at 4–8 °C, (iii) Bring all capsules remaining in the bottle to the next visit, and (iv) avoid consuming probiotic and prebiotic dietary supplements.

*Data Handling and Record Keeping*

Data for screening, enrollment, study progress, and study completion, including demographics, medical history, questionnaires, and adverse reactions were collected during interviews with participants and recorded in the E-platform by RDC Clinical. Radiology data was collected by the radiology site (with body composition data collated by RDC Clinical) and participants' de-identified DXA and QCT data uploaded to Clario according to Best Practices for Dual-Energy X-ray Absorptiometry Measurement and Reporting [1]. All study databases were password protected and backed up on their respective site servers.

*Justification of concentration administered*

For this study, a concentration of $9.5 \times 10^{10}$ CFU/day was chosen according to the following rationale. A completed concentration response study of the SBD111 medical food in mice demonstrated efficacy for the maintenance of BMD at the lumbar spine following estrogen depletion (ovariectomy) with a concentration of $5 \times 10^{8}$ CFU/administration [2]. This concentration allometrically scales to a human equivalent concentration of $1.2 \times 10^{11}$. A previous 28-day toxicity study of the medical food SBD111 in rats at a 20X human safety factor was shown to have no adverse reactions reported [3]. Further, a first in human randomized, double-blind, placebo-controlled safety and tolerability study in 32 healthy adults was completed in 2021, demonstrating no discernable difference in safety or gastrointestinal tolerability between placebo and SBD111 medical food [4].

*Concomitant medications*

Participants taking additional medications for menopausal symptoms or osteoporosis, including but not limited to HT and bisphosphonates, were excluded. Participants consuming probiotics and prebiotics were also excluded. All the medications/supplements which prescribed to participants for other reasons apart from study test article and short-term medication, including antibiotics, were recorded.

*Study product compliance*

In this interventional study, the test product was used for study purposes only. A log was maintained documenting all study test product received and returned to the clinic. For individual participants, accountability was maintained with respect to study test product dispensed, study test product consumed (by counting of the remaining capsules returned at final visit) in online GastroCRF. Once the data was recorded and the bottles/contents were verified by the investigator, the clinical staff disposed of the bottles and product as per Good Clinical Practice.

*Study Outcomes*

Primary outcome measure

The primary outcome of the study was to determine the percent change in BMD at the lumbar spine (2-4 evaluable levels L1-L4) attributable to intervention (relative to placebo), computed at the participant level, between baseline and 12-months of administration amongst participants randomized to the SBD111 medical food or placebo test article.

Secondary outcome measures

The secondary outcomes of the study for participants randomized to active or placebo test article were

i) Additional Bone density outcomes using DXA, QCT, and Bone Turnover markers
   a. Percent change in BMD measured by DXA at the lumbar spine (2-4 evaluable levels L1-L4) between baseline and 6-months
   b. Percent change in trabecular volumetric BMD (vBMD) measured by QCT at the lumbar spine (L1 & L2) between baseline and completion of study period (12 months).
   c. Absolute change in biochemical markers of bone turnover (c-terminal telopeptide (CTX) and procollagen 1 intact N-terminal propeptide (P1NP)) between baseline, 6-months, and completion of study period (12-months).

ii) Inflammatory marker assessments
   a. Absolute change in the circulating marker of inflammation, C-reactive protein (CRP), between baseline, 6-months, and completion of study period (12-months).

Other outcome measures

The other outcomes of this study of participants randomized to intervention or placebo test article were

i) Additional bone density outcomes
   a. Percent change in BMD measured by DXA at the femoral neck and total hip between baseline, 6-months, and completion of study period (12-months).
   b. Percent change in total vBMD (L1-L2) measured by QCT at the lumbar spine between baseline and completion of study period (12 months).
   c. Other characteristics of lumbar spine architecture as measured by QCT between baseline and completion of study period (12 months).

ii) Absolute change in circulating inflammatory cytokines and markers of inflammation (IL-17, TNFα, IL-1β, IL-4, RANKL, IFNγ) between baseline, 6-months, and completion of study period (12 months).

iii) Change in global Menopause Rating Scale (MRS) from baseline to 3, 6, 9, and 12 months.

Exploratory outcome measures

i) Change in gut microbiome composition and function between baseline, 6-months, and completion of study period (12 months)

ii) Absolute change in DXA based lean mass and fat mass from baseline to 6-months, and completion of study period (12 months).

Safety outcome measures

Safety was assessed using the incidence (total count and number of participants experiencing) of adverse events (AE) and serious adverse events (SAE). AE were also characterized according to severity (mild, moderate, severe) and probability of relatedness (unlikely, possible, probable, certain, conditional, unassessible) to trial procedures. Time since test article administration and time to resolution of adverse events was computed. Tolerability was assessed via change in the total Gastrointestinal Tolerability Questionnaire (GITQ) to 12 months.

*Data and sample collection*

*Clinical examination*

Medical history, prescribed and non-prescribed medications, and alcohol intake were recorded during the screening visit. Blood draws and anthropometric measurements were collected using standardized examination procedures and calibrated equipment at four time points (i.e., at enrollment, at baseline, at 26-weeks, and at 52-weeks) (**Table SI1**) [5].

*Blood analysis*

Serum IFNγ, IL-17A, IL-1β, IL-4, IL-6 and TNFα were measured using the Milliplex MAP human high sensitivity T cell magnetic bead kit (Merck; Darmstadt, Germany) as per manufacturer's recommendations. Intra assay variation was 2.7%, 3.0%, 4.5%, 2.7%, 3.3%, and 3.2%, respectively. Interassay variation was 3.5%, 2.5%, 0.3%, 3.9%, 3.2%, and 5.2%, respectively. Serum RANKL was measured using the Milliplex Human RANKL Magnetic bead single plex kit (Merck; Darmstadt, Germany) as per manufacturer's recommendations. Intra assay variation was 2.7%, and interassay variation was 3.0%. All Milliplex assays were analysed on the Merck Magpix instrument. Serum C terminal telopeptides (CTx) and procollagen type 1 N terminal propeptide (P1NP) were measured using the Roche Elecsys β CrossLaps/serum (Roche Diagnostics; Mannheim, Germany) assay and the Elecsys total P1NP assay (Roche

Diagnostics; Mannheim, Germany), respectively. The kits were run on a Cobas e411 analyzer (Roche Diagnostics; Mannheim, Germany), as per manufacturer's recommendations. The intraassay variation was 3.42% (CTx) and 4.37% (P1NP). The interassay variation was 3.41% (CTx) and 4.31% (P1NP). hsCRP was measured in one batch using the Biobase hsCRP test kit, as per manufacturer's recommendations.

*Stool samples*

Stool samples were collected at baseline, at 6-months and at 12-months using a commercially available kit (Microba *Insight*™ Sampling Kit, Microba, Brisbane, Australia) [6]. This kit includes a Coplan FLOQSwab brush in an active drying tube capable of preserving samples at room temperature. The stool kits were delivered to the study site by Microba Pty Ltd (Brisbane, Qld), and kits were provided to participants with instructions for sample collection at home. Participants were instructed to avoid touching the brush and to insert it into the storage tube immediately after sampling. Subsequently, samples collected were sent directly to Microba in pre-paid/pre-labelled envelopes. Samples were then stored at -80°C until study completion and batch analysis. The collection kit also included an instruction booklet for stool sample collection and transportation, gloves, a sterile container, and a sealed plastic pouch.

*Radiology assessment*

The participaiting radiology sites were certified independent laboratories located in Brisbane (QLD) and Sydney (NSW). A dedicated training was provided by Clario to the participating technicians before conducting any radiology testing. BMD of the lumbar spine (L1-L4), and total hip, and femoral neck of the left femur was measured by DXA (Lunar Prodigy, GE Medical Systems, or Hologic Horizon, Hologic, Inc) at baseline, at 6-months, and at 12-months. Quality control and analysis of the DXA scans were performed centrally by blinded technicians (Clario, USA) .

QCT scans of the lumbar spine were performed at baseline and at 12-month timepoints using whole-body CT scanners (Somatom Definition Edge, Siemens, Inc, Incisive, Philips Healthcare, or Lightspeed VCT, GE Medical Systems) via a custom scanning protocol as previously described [7]. Briefly, participants were scanned on top of a bone density calibration phantom from mid-T12 to mid-L3, such that L1 and L2 could be analyzed. The phantom was used to calculate BMD from the measured CT values. For the Siemens scanner a Siemens Syngo Osteo Phantom and for the two other scanners a BDC phantom, (QRM, Germany) was used. Scans were performed at 120kV using 100 mAs and a pitch of 1. CT images were reconstructed with a slice thickness of 1 or 1.25 mm, an reconstruction increment of of 1 or 1.25 mm, a medium sharp kernel and a reconstruction field of view of 150mm that included the vertebra and the calibration phantom.. The long term stability of each CT scanner was monitored via monthly scans of a QRM Spine Phantom (QSP) (QRM, Germany).

All CT scans of a given participant in the trial were conducted using the same CT scanner at each timepoint, and if possible, by the same operator to minimise errors due to machine differences or operator technique differences. As with the DXA data, QCT data were uploaded to Clario for QC and for central analysis by blinded observers using Medical Imaging Analysis Framework (MIAF) Spine software (version 6.0.8R) as previously described [8]. Estimated in vivo root mean square precision errors determined in a group of elderly females 1.2%, 1.5%, and 2.3% for entire vertebral body integral, trabecular, and cortical BMD [9].

*Menopause Symptoms Assessment*

Assessments of menopause symptoms were completed using the Menopause Rating Scale (MRS) at months 3, 6, 9, and 12. The MRS score assesses menopausal symptoms using a scale designed and standardized as a self-administered scale to (a) to assess symptoms/complaints of aging women under different conditions, (b) to evaluate the severity of symptoms over time, and (c) to measure changes pre- and post-menopause replacement therapy [10]. It includes 11 items on a scale from no complaints (0) to very severe symptoms (4). Sub-scores were added to create a composite or total score.

*Safety and Gastrointestinal Tolerability Assessments*

Adverse events are defined as any unfavourable and unintended sign (including an abnormal laboratory finding), symptom, or disease (new or exacerbated) temporally associated with the use of the study product, whether considered related to the medical food under investigation or not. The study investigator monitored each participant for adverse events during the study. All adverse signs or symptoms reported between consent and final follow-up were recorded. Adverse events were reported descriptively by study group and placed in MedDRA categories.

Safety was assessed by incidence of AEs and serious adverse events (SAEs), and tolerability was assessed using the Gastrointestinal Tolerability Questionnaire (GITQ). The gastrointestinal tolerability questionnaire (GITQ) is a 12-item inventory of participant reported gastrointestinal related discomfort. It is a patient self-report questionnaire assessment of the frequency and severity of gastrointestinal symptoms (e.g., gas and abdominal pain), reported on a scale from Mild to Severe for all questions. As probiotics can induce gastrointestinal related symptoms such as gas and bloating, we included a GITQ modified from Pereira et al., 2014 to monitor these potential related symptoms to determine whether the SBD111 medical food causes gastrointestinal discomfort compared to placebo test product comparator [11].

All SAEs, related or not related to the study product, were recorded on electronic case report forms. Serious adverse events were reported in compliance with the requirements of the National Institute of Integrative Medicine Human Research Ethics Committee.

*Microbial whole-genome sequencing*

High-quality genomes were generated for SBD111 strains using a hybrid assembly approach with Oxford Nanopore and Illumina sequencing data. For Illumina sequencing, genomic DNA was extracted from pure cultures using the Zymo Quick-DNA Bacterial/Fungal Kit (Zymo Research). DNA libraries were constructed with the Nextera XT DNA Library Preparation Kit (Illumina Inc.) following the manufacturer's instructions. Library concentration was measured using a Qubit 3.0 Fluorometer (Thermo Fisher Scientific). Sequencing was performed on an Illumina MiSeq platform (MiSeq Control Software v2.6) with 2 × 250 bp paired-end reads. Raw reads were trimmed and filtered based on a Phred score >20 and a minimum fragment length of 50 bp using SolexaQA v3.1.7.1 [12].

For Oxford Nanopore sequencing, genomic DNA from *Pichia kudriavzevii* was extracted using the MasterPure Yeast DNA Purification Kit (Lucigen) and genomic DNA from bacterial strains was extracted using the Quick-DNA Fungal/Bacterial kit (Zymo). DNA libraries were prepared with the Nanopore Genomic DNA by Ligation Kit (SQK-LSK110), and sequencing was performed on the MinION platform (Oxford Nanopore Technologies) using MinKNOW software v20.10.3. Fast5 reads were base called and converted to Fastq format using Guppy v3.2.2 (dna_r9.4.1_450bps_hac.cfg). Read quality was evaluated with NanoFilt v2.7.1, using a Phred score >20 and a minimum fragment length of 1,000 bp [13].

Filtered Nanopore reads were assembled using Flye v1.8, and the assembly was polished with Medaka v0.12.1 (Oxford Nanopore Technologies Ltd.) [14]. Illumina reads were mapped to the assembled contigs for three rounds of polishing using Pilon v1.24 and BWA-MEM v0.7.17 [15,16]. Assembly quality was assessed using QUAST v5.0.2, and genome completeness was evaluated using BUSCO v3 (saccharomycetes_odb10 dataset) for the *P. kudriavzevii* genome and CheckM v1.1.3 for bacterial genomes [17–19].

*Shotgun metagenomic sequencing of stool samples*

Stool sample processing, shotgun metagenomics, and bioinformatics analysis were conducted by Microba (Microba Life Sciences, Australia). DNA was extracted from the stool samples using the DNeasy 96 PowerSoil Pro QIAcube HT Kit (Qiagen), in 2ml deep well plate format as per manufacturer's instructions with a modified initial processing step on the QIAcube HT DNA extraction system (Qiagen). Mechanical lysis was performed with the PowerBead Pro beads (Qiagen) and optimized chemistry enabling more efficient lysis of bacteria and fungi. DNA was quantified using a high sensitivity dsDNA fluorometric assay (QuantIT, ThermoFisher). Samples were required to reach a minimum of 0.2 ng/μL to pass quality control requirements.

DNA sequencing libraries were prepared using the Illumina DNA Prep (M) Tagmentation Kit (Illumina) with IDT for Illumina DNA/RNA UD Index Sets A-D (Illumina) according to manufacturer's instructions. Reaction volume was modified to accommodate processing in a 384-well plate format. Libraries were quantified using a high sensitivity dsDNA fluorometric assay (QuantIT, ThermoFisher), and individual libraries were visualized with capillary gel electrophoresis using the QIAxcel DNA High Resolution Kit (Qiagen). Libraries were pooled at equimolar amounts and the sequencing pool was quantified using a high sensitivity dsDNA fluorometric assay (QuantIT, ThermoFisher) and visualized with capillary gel electrophoresis using the QIAxcel DNA High Resolution Kit (Qiagen). Sequencing was performed on an Illumina NovaSeq6000 instrument following manufacturer's instructions, with v1.5 300 bp PE sequencing reagents to generate 5Gb per sample.

Raw sequencing data was quality-checked, including trimming of adapters and low-quality bases, using Microba's in-house bioinformatics pipeline. Briefly, raw sequencing data was demultiplexed and adaptor trimmed using Illumina BaseSpace Bcl2fastq2 (v2.20) allowing one mismatch in index sequences. Raw reads were then quality trimmed and residual adaptors removed using Trimmomatic v0.36 with the following parameters: -phred33 LEADING:3 TRAILING:3 [20]. Human DNA was identified and removed by aligning reads to the human genome reference assembly 38 (GRCh38.p12, GCF_000001405) using bwa-mem v0.7.18 with default parameters except minimum seed length set to -k 31 [16]. Alignments were further filtered using SAMtools v1.7, with flags -ubh -f1 -F2304 [21].

*Stool metagenome analysis*

Taxonomic profiles of stool metagenomes were generated using the Microba Community Profiler (MCP) v3.1 ([11]) with the Microba Genome Database (MGDB) v4.1 as the reference genome database [22]. The MGDB is an expanded version of the GenBank, National Institute of Health (US) and the Genome Taxonomic Database (GTDB) of the Australian Centre for Ecogenomics at its core. This analysis estimates the relative abundance of bacterial, archael, and eukaryotic community members, as well as the bacterial diversity within a sample (alpha diversity) and between all the samples (beta diversity). Sequencing reads were mapped to genomes within the MGDB, and relative abundances of species clusters were estimated and reported. Metabolic pathway and gene abundance quantification was performed using the Microba Feature Counter (MFC) v1.6 with the MGDB as the reference database. In the first step, MCP-derived read mappings aligning to annotated gene sequences of a length equal to or greater than a configurable threshold (e.g., 10 base pairs) were counted. In the second step, Enzyme Commission (EC) annotations were used to assess MetaCyc pathway completeness, applying a threshold of >80% [23]. Pathway abundance per species was reported by averaging read counts across all genes within each pathway.

Bacterial species diversity analysis was conducted using rarefied count data standardized to 9 million reads per sample. Alpha diversity metrics, including Richness and Shannon Diversity Index, were calculated. For beta diversity, Bray-Curtis dissimilarities were computed and visualized using Principal Coordinates Analysis (PCoA).

*SBD111 strain detection in stool metagenomes*

Detection of SBD111 strains in stool metagenomes was performed using Strainer2 and custom bioinformatics scripts [24]. Briefly, to identify strain-specific genomic sequences, SBD111 strain genomes were fragmented into overlapping 31 bp k-mers and then mapped to a custom database of human stool metagenomic samples, reference genomes closely related to the product strains (Average Nucleotide Identity >98%), and the MetaPhlAn4 database (mpa_vJan21_CHOCOPhlAnSGB_202103) [25]. K-mer frequencies were calculated across the custom database, and rare k-mers were selected for each SBD111 genome based on specific thresholds, to represent strain-specific sequences: for *L. plantarum* SBS04260, *L. brevis* SBS04254, and *Leuc. mesenteroides* SBS02455, the 1% of k-mers least commonly detected in the training database were selected as strain-specific k-mers, and for *P. kudriavzevii* SBS04263, the 23% of k-mers least commonly detected in the training database were selected as strain-specific k-mers.

For each SBD111 component strain, strain-specific k-mers were then mapped to participant stool metagenomes to calculate k-mer coverage and depth. Coverage was defined as the proportion of unique k-mers observed in a metagenome sample relative to the total k-mers present in the strain-specific k-mer set. K-mer depth was calculated as the total number of observed k-mers divided by the total k-mers present in the strain-specific k-mer set. Strain relative abundance was estimated by multiplying k-mer depth by the strain's genome size and dividing by the metagenome size. A strain was considered detected in a sample when the k-mer coverage exceeded 0.03x for bacterial strains or 0.01x for fungal strains. These coverage thresholds were selected to eliminate false positive detection in training metagenome samples.

*Microbiome statistical analysis*

Statistical analysis was performed for evaluating differences in relative abundance of bacterial taxa and functional pathways. For paired comparisons between two samples at a single time point (e.g., baseline comparisons), Welch's paired two-sample t-test was used to assess if the value of outcome variable measure differs between the two repeated measures study groups of interest. Linear mixed effect models were used to assess change across repeated measure variables between study groups of interest. The models included random effects for the units that were measured repeatedly, e.g. participant, and fixed effects for the repeated measures variable, study group, and interactions between the repeated measures variable and study group (e.g., study group x time point). P values were generated by nested model test of the significance of including the corresponding fixed effect. For all statistical tests, the Benjamini–Hochberg false discovery rate adjustment was used to correct for multiple hypothesis testing [26].

*Withdrawals and Dropouts*

A participant could have been withdrawn from the study if: 1) the participant withdrew informed consent (with or without explanation, including those lost to follow-up); 2) AE(s) or SAE(s) (including pathology abnormalities) occured which, according to the investigator, made study continuation not possible; 3) a concomitant condition was identified in which the prescribed additional therapy was prohibited by protocol as per the participant selection criteria; 4) protocol deviation, affecting the study results occured; 5) discretion of the investigator for any reason, if it was felt that further continuation in the study would adversely affect the participant, or, in the interests of the study.

Participant withdrawls were documented, and all SAEs were reported to the investigators. If any participant had been enrolled for study participation and the participant decided to withdraw from the study at any point time prior to study completion, all study procedures applicable at final assessment interview were performed at the time of study withdrawal so long as the participant was agreeable. If the participant failed to complete the final visit assessments, they were included in the modified intention-to-treat population analysis and not the per protocol population.

*Ethical considerations*

Ethics oversight was sought from and approved by the National Institute of Integrative Medicine (NIIM) and ratified by additional radiology site ethics committees, in accordance with ICH Good Clinical Practice Guidelines. Protocol amendments were only made after consultation with trial management committee, and all protocol changes were approved by the institutional Human Research Ethics Committees (HREC) prior to implementation. Tolerability and AEs were recorded at regular intervals and reviewed at 3 monthly intervals by an independent medical practitioner. Assessment of risk of osteopenia continued after enrollment in the trial. Bone density assessments at month 6 were flagged if results that show greater than 7% loss of bone density from the baseline assessment, and participants were referred by the medical practitioner/specialist for them to obtain medical advice before continuing in the study.

*Ethics and Dissemination*

The study was conducted according to the Note for Guidance on Good Clinical Practice (CPMP/ICH/135/95) annotated with Therapeutic Goods Administration Drug Safety and Evaluation Branch comments (July 2000) and in compliance with applicable Australian laws and regulations. The study was performed in accordance with the NHMRC Statement on Ethical Conduct in Research Involving Humans 2007 (updated May 2015), the NHMRC Australian Code for the Responsible Conduct of Research 2007, and the principles laid down by the World Medical Assembly in the Declaration of Helsinki 2008.

**Supplemental Results**

*SBD111 administration did not alter menopause symptom severity*

To evaluate the effects of twice daily administration of SBD111 on symptoms of menopause, study participants were asked to complete an 11-item Menopause Rating Scale (MRS) detailing the severity of physical and psychological menopause symptoms at baseline and at 3, 6, 9, and 12-months. Analysis of the resulting data revealed no significant effects of SBD111 on overall MRS scores or on the somatic, psychological, or urogenital components of the survey at any timepoint evaluated (**Table SI9**). These observations suggest that twice daily SBD111 administration does not improve or worsen menopause symptoms in early postmenopausal women.

*Effects of SBD111 administration on the gut microbiota*

To characterize the effects of twice daily administration of SBD111 on the structure and function of the gut microbiota of early postmenopausal women, stool samples collected at baseline, 6-months, and 12-months were subjected to metagenomic DNA sequencing. We first aimed to detect and determine the abundance of SBD111 component strains in participants' stool metagenomes using custom designed sets of strain-specific k-mer sequences for each SBD111 strain (see **Supplemental Methods**). This analysis confirmed that no SBD111 component strains were present in any stool samples collected from participants in the placebo group. Coverage of SBD111 strain genomes was also absent from most baseline samples in the SBD111 intervention group but robustly detected at 6- and 12-months (**Fig SI2a**). Strains of *L. plantarum*, *L. brevis*, and *Leuc. mesenteroides* specific to the SBD111 medical food were detected in two baseline samples from participants in the SBD111 group. Given the absence of these strains in all placebo group samples (n = 321) and that stool samples kits were provided for at-home collection concomitantly with study product, these baseline stool samples may have been collected by participants after the initial administration of SBD111. This was confirmed for one such participant, and we hypothesize this is the case for the remaining subject. SBD111 strains of *L. plantarum* SBS04260, *L. brevis* SBS04254, and *Leuc. mesenteroides* SBS02455 were detected in >70% of subjects in the SBD111 intervention group at 6- and 12-months (**Fig SI2b**). By contrast, *P. kudriavzevii* SBS04263 was detected in 43% and 36% of participants in the SBD111 group at 6- and 12-months, respectively (**Fig SI2b**). The reduced rate of detection for *P. kudriavzevii* SBS04263 may be due to the lower concentration of this strain in the SBD111 medical food (**Table SI2**). Overall, these findings confirm the presence of SBD111 strains in the stool metagenomes of subjects receiving the medical food at 6- and 12-months as well as the absence of these strains in the stool metagenomes of subjects that received placebo.

We next sought to evaluate whether administration of the SBD111 synbiotic medical food altered the representation of microbial species and functional pathways in the stool metagenomes of early postmenopausal subjects. Linear mixed effects regression (LMER) models using paired stool metagenome data from baseline and 12 months revealed no significant differences in the number of bacterial species detected or Shannon indices, a measure of community diversity, associated with product administration ($p > 0.05$, LMER interaction term; **Fig SI2c-d**). By contrast, administration of SBD111 was associated with significantly increased richness of MetaCyc functional pathways at 12-months compared to baseline ($p = 0.03$, LMER interaction term), though this difference was not observed in Shannon indices of functional pathways represented (**Fig SI2e-f**). These observations indicate that while SBD111 did not alter the diversity of the gut microbial community at a species level at 12 months, administration of the synbiotic medical food modestly increased the functional diversity in this community.

We then evaluated changes in these stool metagenomes at the level of individual species and functional pathways using LMER models. Consistent with strain detection analysis described above, this analysis confirmed significantly increased abundance of *L. plantarum*, *L. brevis*, *Leuc. mesenteroides*, and *P. kudriavzevii* in the stool metagenomes of subjects in the SBD111 group at 6- and 12-months relative to baseline and to placebo ($p<0.001$, LMER interaction term with FDR correction; **Fig SI3a-h**). In addition to the species present in SBD111 synbiotic medical food, administration of SBD111 was also associated with significantly decreased abundance of a *Clostridium* strain CAG-269 sp900556695 at 12-months ($p<0.001$, LMER interaction term with FDR correction). Furthermore, LMER analysis identified two MetaCyc functional pathways with significantly increased abundance at 6- and 12-months in SBD111 recipients as compared to placebo: (i) anthranilate degradation IV and (ii) trehalose degradation III ($p<0.001$; **Fig S3i-l**). The anthranilate degradation IV pathway is present in the genome of *L. plantarum* SBS04260, and the trehalose degradation II pathway is present in the genomes of *L. plantarum* SBS04260*, L. brevis* SBS04254*,* and *Leuc. mesenteroides* SBS02455. As such, enrichment of these in the stool metagenomes of SBD111 recipients is consistent with increased abundance of these strains. These data confirm that while SBD111 administration does not substantially alter the species composition of the gut microbiome from postmenopausal women, this does result in increased abundance of SBD111 strains as well as pathways represented in these strains' genomes.

The effects of SBD111 administration on the metagenomes of subjects with osteopenia at baseline were similar to our observations of the full study cohort. In this group, no significant effects were observed on species or MetaCyc pathway richness or Shannon indices at 6- or 12-months. However, administration of SBD111 was associated with significant enrichment of *L. plantarum*, *L. brevis*, *Leuc. mesenteroides*, and *P. kudriavzevii* species as well as the anthranilate degradation IV and trehalose degradation III MetaCyc pathways at 6- and 12-months relative to baseline and to placebo in subjects with osteopenia at baseline ($p<0.001$, LMER interaction term with FDR correction).

*Supplemental Information Figures*

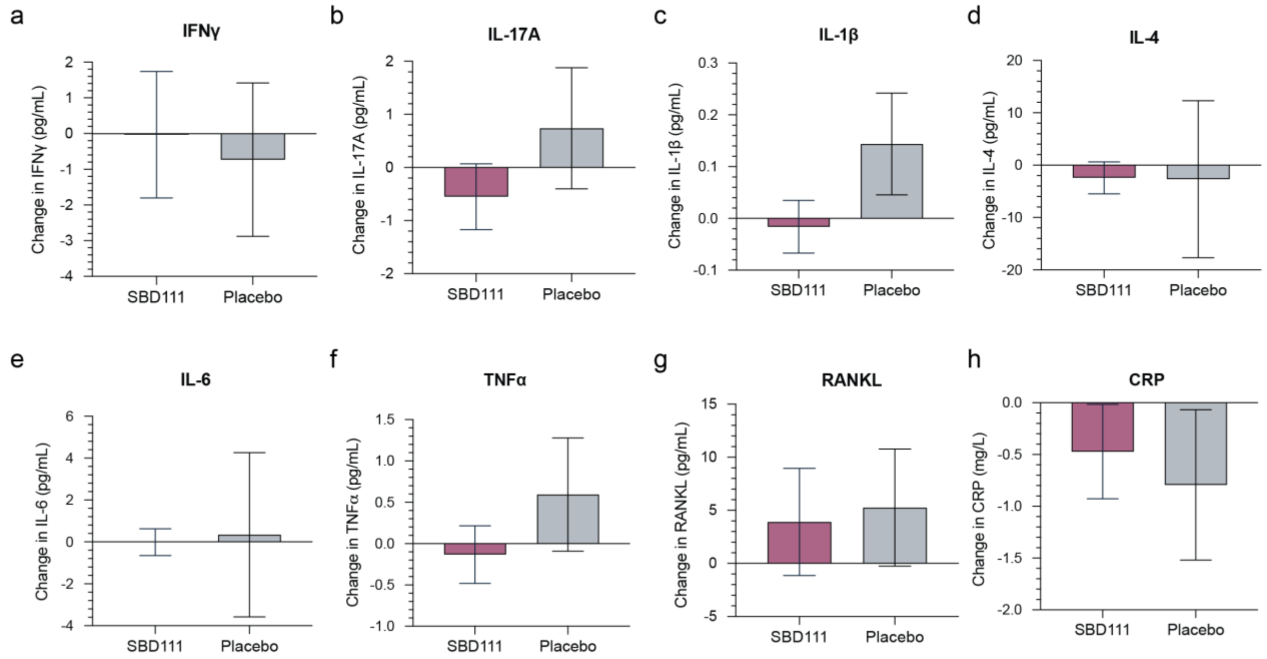

**Fig SI1** Change in concentration of serum (a) IFNy, (b) IL-17A, (c) IL-1b, (d) IL-4, (e) IL-6, (f) TNFa, (g) RANKL, and (h) CRP from baseline to 12 months for subjects randomized to receive either twice daily SBD111 or maltodextrin placebo. Values represent mean ± standard error of the mean concentration change for each group. Positive values indicate increased concentrations from baseline to 12 months

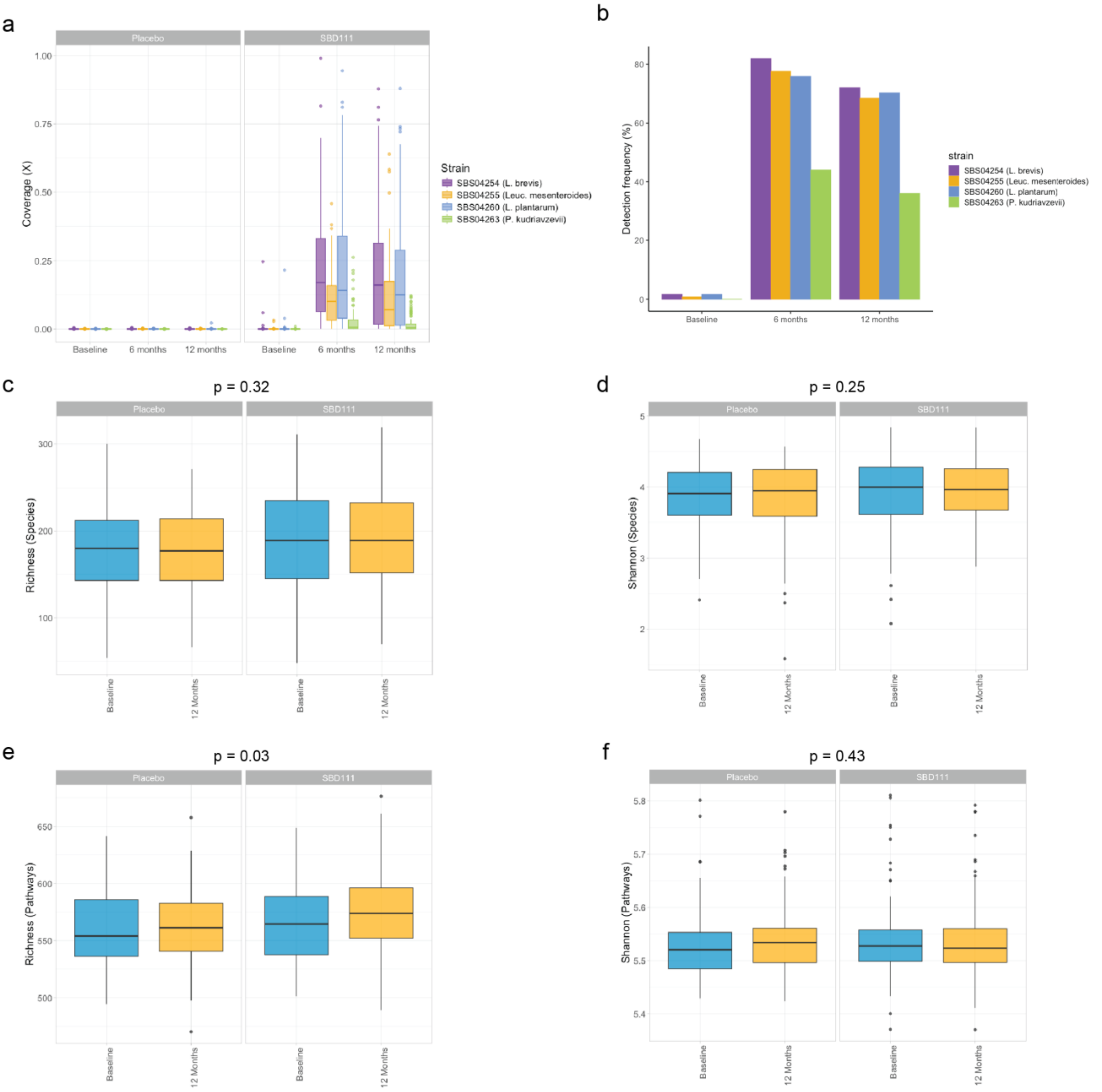

**Fig SI2** (a) Fold-coverage of genome-specific k-mer sets and (b) detection frequency in stool metagenomes for each microbial strain comprising SBD111 (*L. plantarum* SBS04260, *L. brevis* SBS04254, *Leuc. mesenteroides* SBS02455, and *P. kudriavzevii* SBS04263) at baseline, 6 months, and 12 months for subjects randomized to receive twice daily SBD111 or maltodextrin placebo. For (a), box plots depict median, IQR, and 95% CI, with outliers. Each point represents an individual stool metagenome sample. (c-f) Box plots comparing (c) bacterial species richness, (d) Shannon diversity indices for bacterial species, (e) MetaCyc functional pathway richness, and (f) MetaCyc functional pathway Shannon diversity indices at baseline and 12 months for subjects randomized to receive twice daily SBD111 or maltodextrin placebo. Lines indicate paired stool metagenome samples for individual subjects. For (c-f), box plots depict median, IQR, and 95% CI. P-values represent significance estimated from linear mixed effects regression model interaction terms after FDR correction

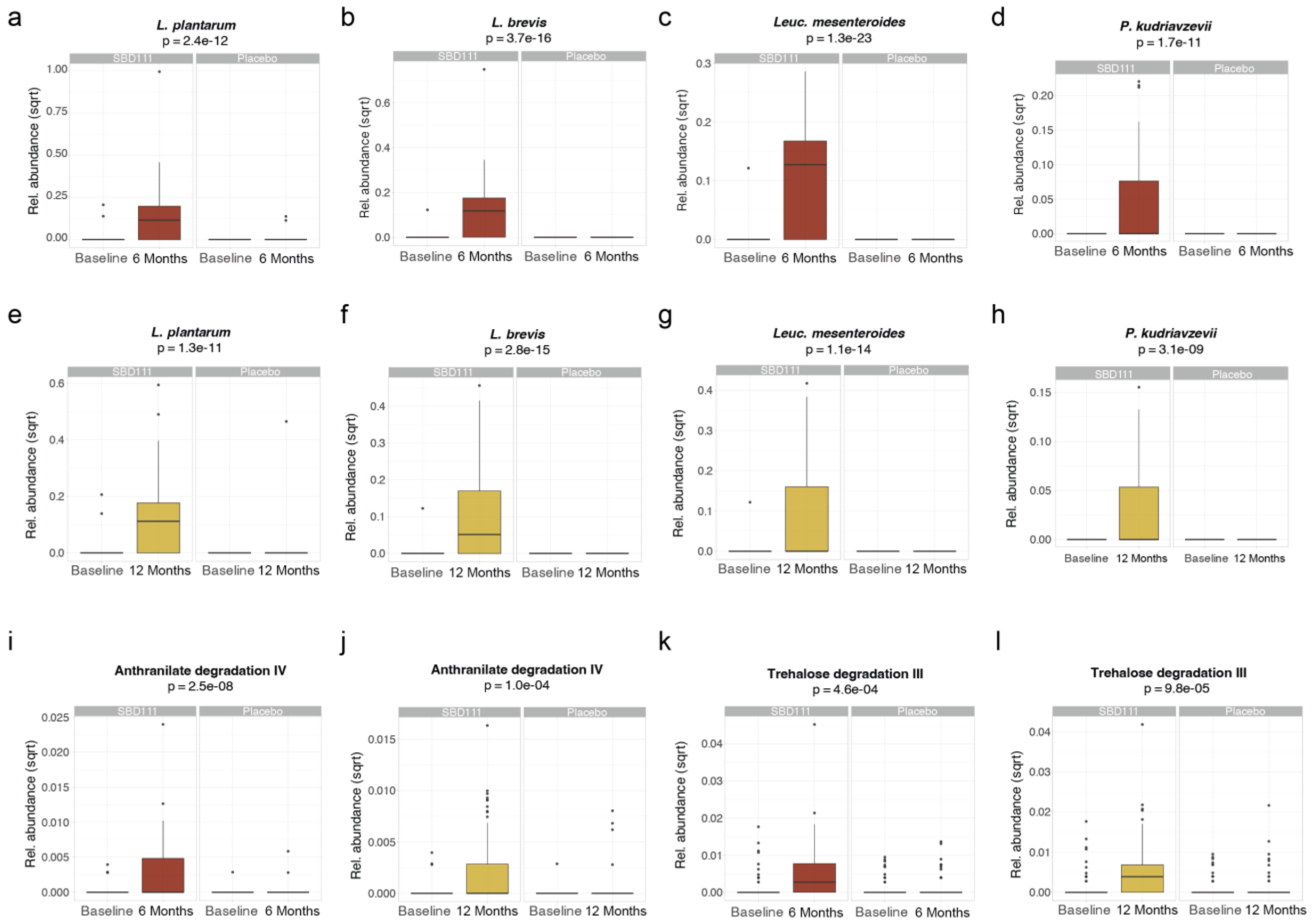

**Fig SI3** (a-d) Box plots comparing relative abundance of (a) *L. plantarum*, (b) *L. brevis*, (c) *Leuc. mesenteroides*, and (d) *P. kudriavzevii* at baseline and 6 months for subjects randomized to receive twice daily SBD111 or maltodextrin placebo. (e-h) Box plots comparing relative abundance of (e) *L. plantarum*, (f) *L. brevis*, (g) *Leuc. mesenteroides*, (h) *P. kudriavzevii* at baseline and 12 months for subjects randomized to receive twice daily SBD111 or maltodextrin placebo. (i-j) Box plots comparing relative abundance of anthranilate degradation IV at (i) baseline and 6 months or (j) baseline and 12 months for subjects randomized to receive twice daily SBD111 or maltodextrin placebo. (k-l) Box plots comparing relative abundance of trehalose degradation III at (k) baseline and 6 months or (l) baseline and 12 months for subjects randomized to receive twice daily SBD111 or maltodextrin placebo. For (a-l), box plots depict median, IQR, and 95% CI. P-values represent significance estimated from linear mixed effects regression model interaction terms after FDR correction

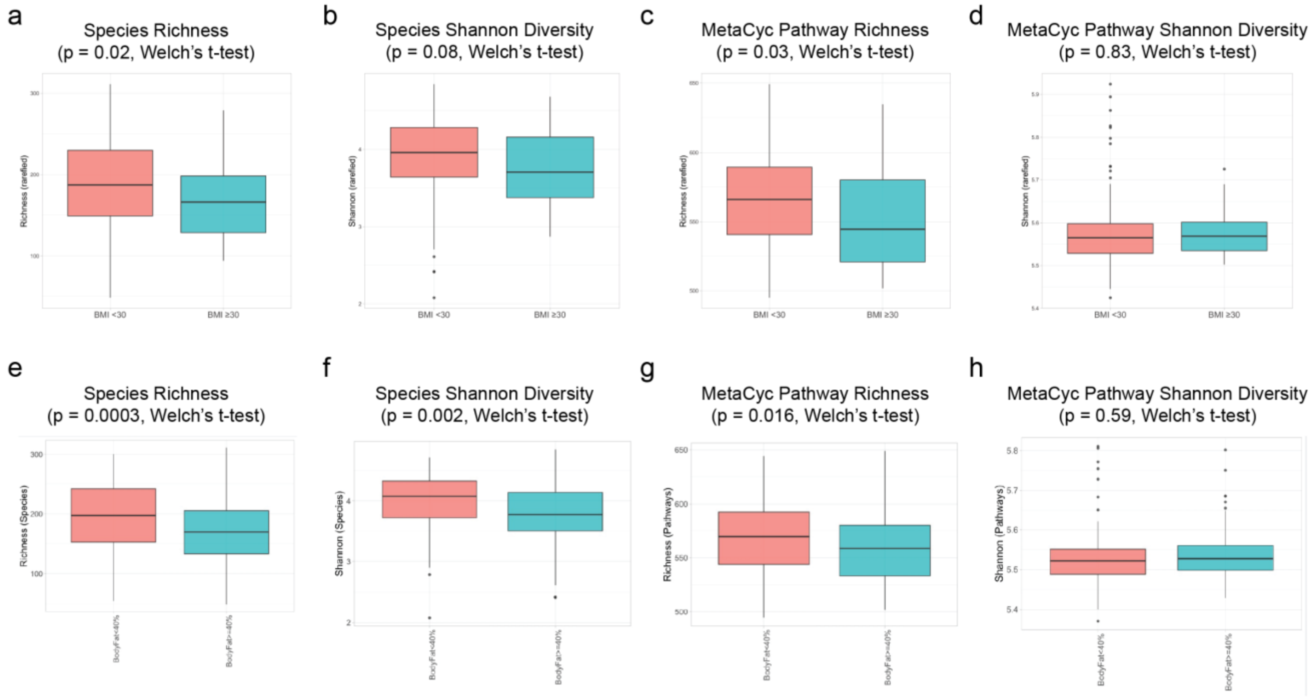

**Fig SI4** (a-d) Box plots comparing (a) bacterial species richness, (b) Shannon diversity indices for bacterial species, (c) MetaCyc functional pathway richness, and (d) MetaCyc functional Shannon diversity indices at baseline for subjects with baseline BMI ³ 30 or BMI < 30. For (a-d), box plots depict median, IQR, and 95% CI. Points represent individual stool metagenome samples. P-values represent Welch's t-tests

***Supplemental Information Tables***

**Table SI1** Schedule of enrollment, interventions, and assessments

| | **STUDY PERIOD** | | | | | | | | | | |
|---|---|---|---|---|---|---|---|---|---|---|---|
| | **Enrollment** | **Allocation** | **Time post-allocation (weeks)** | | | | | | | | **Close-out** |
| | | **Baseline** | | | | | | | | | |
| **TIMEPOINT (weeks)** | **0** | **0** | $t_1$ | $t_2$ | $t_4$ | $t_8$ | $t_{16}$ | $t_{24}$ | $t_{32}$ | $t_{40}$ | $T_{52}$ |
| **ENROLLMENT:** | | | | | | | | | | | |
| Eligibility screen | X | | | | | | | | | | |
| Informed consent | X | | | | | | | | | | |
| Demographics<br>Vital Signs | X | X | | | | | | X | | | X |
| Blood draw | X | X | | | | | | X | | | X |
| DXA scan | X | | | | | | | X | | | X |
| qCT Scan | | X | | | | | | | | | X |
| Stool sample | | X | | | | | | X | | | X |
| QoL | | X | | | | X | X | | X | | X |
| FFQ and Exercise log | | X | | | | | | X | | | X |
| Allocation | | X | | | | | | | | | |
| **INTERVENTIONS:** | | | | | | | | | | | |
| *Test Article A (SBD111)*<br>*Adjunctive Vitamin $D_3$* | | | | | | | | | | | |
| *Test Article B (Placebo)*<br>*Adjunctive Vitamin $D_3$* | | | | | | | | | | | |
| **ASSESSMENTS:** | | | | | | | | | | | |
| *Tolerability Assessment* | | X | X | X | X | X | X | X | X | X | X |
| *Safety Assessment* | | | X | X | X | X | X | X | X | X | X |
| *Compliance* | | | | | | X | X | X | X | X | X |

**Table SI2** Composition of SBD111 Medical Food

| **Ingredient** | **Amount per Capsule (CFU)*** | **Daily Amount in Four Capsules (CFU)** | **Ingredient in Each Capsule (mg)** |
|---|---|---|---|
| *Levilactobacillus brevis* SBS04254 | $7.5x10^{9}$ | $3.0x10^{10}$ | 14-24 |
| *Lactiplantibacillus plantarum* SBS04260 | $7.5x10^{9}$ | $3.0x10^{10}$ | 10-14 |
| *Leuconostoc mesenteroides* SBS02455 | $7.5x10^{9}$ | $3.0x10^{10}$ | 13-23 |
| *Pichia kudriavzevii* SBS04263 | $1.25x10^{9}$ | $5.0x10^{9}$ | 75-100 |
| Oligofructose | NA | NA | 130-180 |
| Dried ground blueberry powder | NA | NA | 130-180 |
| Magnesium stearate | NA | NA | 5-10 |
| Silicon dioxide | NA | NA | 5-10 |
| **Total** | **$2.375x10^{10}$** | **$9.5x10^{10}$** | **440-490** |

*CFU = colony forming units; NA = Not Applicable, not part of microorganism dose

**Table SI3** Summary of adverse events (AEs)

| **Number of AEs per participant** | **Mean** | **SD** | **Pearson's Chi$^2$ p-value** |
|---|---|---|---|
| SBD111 | 2.48 | 1.55 | 0.6 |
| Placebo | 2.63 | 1.71 | |
| **Relatedness to study product** | **Possibly related AEs** | **Unrelated AEs** | **Pearson's Chi$^2$ p-value** |
| SBD111 | 46 (70%) | 20 (30%) | 0.87 |
| Placebo | 41 (68%) | 19 (32%) | |

**Table SI4** qCT percent change in lumbar spine (0 - 12 months) for study cohort and prespecified subgroups

| Measurement | N | Mean (95% CI) | p-val | Measurement | N | Mean (95% CI) | p-val |
|---|---|---|---|---|---|---|---|
| **Trabecular vBMD, %** | | | | **Trabecular BMC, %** | | | |
| All Subjects | 208 | -0.86 (-2.16, 0.44) | 0.90 | All Subjects | 208 | -1.61 (-3.41, 0.20) | 0.96 |
| BMI < 30 | 170 | -0.79 (-2.20, 0.62) | 0.86 | BMI < 30 | 170 | -1.08 (-3.08, 0.93) | 0.86 |
| BMI ≥ 30 | 38 | -1.30 (-4.89, 2.29) | 0.77 | BMI ≥ 30 | 38 | -4.02 (-8.55, 0.52) | 0.96 |
| Osteopenic | 78 | -0.54 (-2.77, 1.69) | 0.68 | Osteopenic | 78 | -0.78 (-3.28, 1.73) | 0.73 |
| Non-osteopenic | 130 | -1.05 (-2.67, 0.58) | 0.90 | Non-osteopenic | 130 | -2.12 (-4.61, 0.38) | 0.95 |
| < 4 years in menopause | 123 | -0.61 (-2.41, 1.20) | 0.75 | < 4 years in menopause | 123 | -1.24 (-3.85, 1.37) | 0.83 |
| ≥ 4 years in menopause | 83 | -1.23 (-3.00, 0.54) | 0.91 | ≥ 4 years in menopause | 83 | -2.18 (-4.59, 0.23) | 0.96 |
| **Total vBMD, %** | | | | **Total BMC, %** | | | |
| All Subjects | 208 | -0.29 (-1.16, 0.58) | 0.75 | All Subjects | 208 | -0.31 (-1.32, 0.70) | 0.73 |
| BMI < 30 | 170 | -0.43 (-1.39, 0.53) | 0.81 | BMI < 30 | 170 | -0.50 (-1.60, 0.59) | 0.82 |
| BMI ≥ 30 | 38 | 0.15 (-2.08, 2.39) | 0.44 | BMI ≥ 30 | 38 | 0.28 (-2.47, 3.02) | 0.42 |
| Osteopenic | 78 | 0.03 (-1.50, 1.55) | 0.49 | Osteopenic | 78 | -0.31 (-2.14, 1.52) | 0.63 |
| Non-osteopenic | 130 | -0.49 (-1.55, 0.58) | 0.82 | Non-osteopenic | 130 | -0.32 (-1.52, 0.88) | 0.70 |
| < 4 years in menopause | 123 | -0.24 (-1.44, 0.95) | 0.66 | < 4 years in menopause | 123 | 0.10 (-1.23, 1.43) | 0.44 |
| ≥ 4 years in menopause | 83 | -0.35 (-1.51, 0.81) | 0.72 | ≥ 4 years in menopause | 83 | -0.93 (-2.47, 0.62) | 0.88 |
| **Cortical vBMD, %** | | | | **Cortical BMC, %** | | | |
| All Subjects | 208 | -0.15 (-1.20, 0.91) | 0.61 | All Subjects | 208 | 0.64 (-1.84, 3.12) | 0.31 |
| BMI < 30 | 170 | 0.01 (-1.16, 1.19) | 0.49 | BMI < 30 | 170 | -0.60 (-3.16, 1.95) | 0.68 |
| BMI ≥ 30 | 38 | -0.81 (-3.37, 1.76) | 0.74 | BMI ≥ 30 | 38 | 5.38 (-2.23, 13.00) | 0.08 |
| Osteopenic | 78 | 0.90 (-0.81, 2.62) | 0.15 | Osteopenic | 78 | 0.18 (-3.54, 3.90) | 0.46 |
| Non-osteopenic | 130 | -0.78 (-2.12, 0.57) | 0.87 | Non-osteopenic | 130 | 0.90 (-2.42, 4.21) | 0.30 |
| < 4 years in menopause | 123 | -0.59 (-1.92, 0.73) | 0.81 | < 4 years in menopause | 123 | 0.67 (-2.57, 3.92) | 0.34 |

| Measurement | N | Mean (95% CI) | p-val | Measurement | N | Mean (95% CI) | p-val |
|---|---|---|---|---|---|---|---|
| ≥ 4 years in menopause | 83 | 0.62 (-1.10, 2.33) | 0.24 | ≥ 4 years in menopause | 83 | 0.56 (-3.42, 4.55) | 0.39 |
| **Subcortical vBMD, %** | | | | **Subcortical BMC, %** | | | |
| All Subjects | 208 | -0.96 (-2.77, 0.85) | 0.85 | All Subjects | 208 | -1.04 (-3.34, 1.25) | 0.81 |
| BMI < 30 | 170 | -0.29 (-2.27, 1.70) | 0.61 | BMI < 30 | 170 | -0.11 (-2.66, 2.44) | 0.53 |
| BMI ≥ 30 | 38 | -3.99 (-8.57, 0.58) | 0.96 | BMI ≥ 30 | 38 | -5.23 (-10.83, 0.36) | 0.97 |
| Osteopenic | 78 | -0.49 (-2.92, 1.95) | 0.65 | Osteopenic | 78 | -0.46 (-3.21, 2.30) | 0.63 |
| Non-osteopenic | 130 | -1.24 (-3.77, 1.28) | 0.83 | Non-osteopenic | 130 | -1.40 (-4.72, 1.92) | 0.80 |
| < 4 years in menopause | 123 | -0.73 (-3.36, 1.90) | 0.71 | < 4 years in menopause | 123 | -0.74 (-4.15, 2.67) | 0.67 |
| ≥ 4 years in menopause | 83 | -1.23 (-3.60, 1.14) | 0.85 | ≥ 4 years in menopause | 83 | -1.44 (-4.31, 1.42) | 0.84 |

vBMD: Volumetric Bone Mineral Density; BMC: Bone Mineral Content. Values represent mean effect and 95% confidence interval in percent change of BMD. Positive values correspond to decreased bone loss in SBD111 recipients. P-values represent analysis by one-sided Student's t-test, pooled variance.

**Table SI5** Change in serum biomarkers of bone turnover

| | N | Month 12 Change from Baseline Mean (SD) | | p-val | | N | Month 6 Change from Baseline Mean (SD) | | p-val |
|---|---|---|---|---|---|---|---|---|---|
| | | **SBD111** | **Placebo** | | | | **SBD111** | **Placebo** | |
| **Serum P1NP (ng/mL)** | | | | | | | | | |
| All Subjects | 217 | -3.290 (19.011) | -2.709 (16.290) | 0.810 | | 211 | 0.711 (18.414) | -0.623 (18.809) | 0.603 |
| BMI ≥ 30 | 41 | -3.365 (13.328) | 0.418 (12.922) | 0.375 | | 40 | -0.937 (16.960) | 4.908 (28.901) | 0.427 |
| Body Fat ≥ 40% | 101 | -2.988 (15.914) | -3.749 (17.639) | 0.885 | | 97 | -0.431 (14.507) | -0.202 (21.415) | 0.953 |
| Osteopenic | 78 | -5.886 (19.577) | -5.450 (15.499) | 0.916 | | 78 | -1.699 (16.426) | -1.958 (22.046) | 0.953 |
| **Serum CTX (ng/mL)** | | | | | | | | | |
| All Subjects | 217 | -0.009 (0.139) | -0.002 (0.134) | 0.695 | | 211 | 0.015 (0.110) | 0.018 (0.127) | 0.874 |
| BMI ≥ 30 | 41 | **-0.034 (0.137)** | **0.048 (0.107)** | **0.049** | | 40 | -0.021 (0.118) | 0.029 (0.152) | 0.254 |
| Body Fat ≥ 40% | 101 | -0.026 (0.142) | -0.002 (0.132) | 0.379 | | 97 | -0.015 (0.102) | 0.015 (0.141) | 0.244 |
| Osteopenic | 78 | 0.003 (0.121) | -0.015 (0.114) | 0.510 | | 78 | 0.024 (0.096) | 0.012 (0.157) | 0.675 |

**Table SI6** Bacterial species with differential abundance in stool metagenomes of subjects with BMI ≥ 30 compared to those with BMI < 30 at baseline

| **Species** | **Abundance Subjects with BMI < 30 at Baseline** | | **Abundance Subjects with BMI ≥ 30 at Baseline** | | **Fold Change Log2(BMI ≥ 30 / BMI < 30)** | **p-value (Welch's t-test)** | **p-value (Welch's t-test with FDR correction)** |
|---|---|---|---|---|---|---|---|
| | Mean | SD | Mean | SD | | | |
| CAG-170 sp900549635 | 0.025 | 0.060 | 0.003 | 0.010 | -3.2 | 5.00E-07 | 2.50E-04 |
| Faecimorpha stercoravium | 0.004 | 0.012 | 0.000 | 0.000 | na | 6.40E-07 | 2.50E-04 |
| Clostridium sp000435835 | 0.040 | 0.095 | 0.005 | 0.016 | -2.9 | 8.80E-06 | 1.70E-03 |
| Eubacterium_I sp900557275 | 0.005 | 0.016 | 0.000 | 0.000 | na | 6.40E-06 | 1.70E-03 |
| Anaerostipes hadrus A | 0.140 | 0.420 | 0.011 | 0.041 | -3.7 | 1.40E-05 | 2.20E-03 |
| CAKTXU01 MIC52682 | 0.003 | 0.016 | 0.000 | 0.000 | na | 7.30E-05 | 9.60E-03 |
| Scatenecus faecavium | 0.010 | 0.040 | 0.000 | 0.000 | na | 1.10E-04 | 1.20E-02 |
| Dysosmobacter sp900542115 | 0.003 | 0.012 | 0.000 | 0.000 | na | 1.80E-04 | 1.80E-02 |
| Butyricicoccus_A intestinisimiae | 0.030 | 0.110 | 0.002 | 0.013 | -3.9 | 6.50E-04 | 3.90E-02 |
| Butyrivibrio_A crossotus | 0.190 | 0.950 | 0.000 | 0.000 | na | 5.30E-04 | 3.90E-02 |
| CAG-273 sp905214245 | 0.006 | 0.027 | 0.000 | 0.000 | na | 5.40E-04 | 3.90E-02 |
| CAG-353 sp900066885 | 0.089 | 0.430 | 0.000 | 0.000 | na | 5.70E-04 | 3.90E-02 |
| Faecimorpha MIC46038 | 0.003 | 0.017 | 0.000 | 0.000 | na | 6.30E-04 | 3.90E-02 |
| CAG-269 sp934177555 | 0.002 | 0.009 | 0.000 | 0.000 | na | 7.20E-04 | 4.00E-02 |
| CAG-302 sp000431795 | 0.062 | 0.190 | 0.013 | 0.056 | -2.3 | 1.20E-03 | 4.30E-02 |
| Catenibacillus sp018369015 | 0.004 | 0.016 | 0.001 | 0.003 | -3.1 | 9.20E-04 | 4.30E-02 |
| Clostridium sp900539375 | 0.034 | 0.200 | 0.000 | 0.000 | na | 1.10E-03 | 4.30E-02 |
| Dysosmobacter sp905200545 | 0.009 | 0.045 | 0.000 | 0.000 | na | 1.00E-03 | 4.30E-02 |
| Eubacterium_R sp000434995 | 0.110 | 0.310 | 0.019 | 0.110 | -2.5 | 1.20E-03 | 4.30E-02 |
| Massilistercora timonensis | 0.010 | 0.054 | 0.000 | 0.000 | na | 9.60E-04 | 4.30E-02 |
| Prevotella timonensis | 0.011 | 0.052 | 0.000 | 0.000 | na | 1.10E-03 | 4.30E-02 |
| RGIG3159 MIC50741 | 0.001 | 0.007 | 0.000 | 0.000 | na | 1.10E-03 | 4.30E-02 |

| Species | Abundance Subjects with BMI < 30 at Baseline | | Abundance Subjects with BMI ≥ 30 at Baseline | | Fold Change Log2(BMI ≥ 30 / BMI < 30) | p-value (Welch's t-test) | p-value (Welch's t-test with FDR correction) |
|---|---|---|---|---|---|---|---|
| | Mean | SD | Mean | SD | | | |
| Anaerotardibacter sp900538545 | 0.007 | 0.036 | 0.000 | 0.000 | na | 1.40E-03 | 4.40E-02 |
| UBA11524 sp000437595 | 0.690 | 1.000 | 0.290 | 0.620 | -1.3 | 1.40E-03 | 4.40E-02 |
| UMGS1781 sp900553695 | 0.010 | 0.046 | 0.000 | 0.000 | na | 1.30E-03 | 4.40E-02 |
| Merdiplasma excrementigallinarum | 0.006 | 0.031 | 0.000 | 0.000 | na | 1.50E-03 | 4.50E-02 |
| Faecaligallichristensenella faecipullorum | 0.003 | 0.014 | 0.000 | 0.000 | na | 1.70E-03 | 4.70E-02 |
| Fumia MIC53095 | 0.001 | 0.005 | 0.000 | 0.000 | na | 1.70E-03 | 4.70E-02 |
| TWA4 MIC45161 | 0.006 | 0.032 | 0.000 | 0.000 | na | 1.60E-03 | 4.70E-02 |

na: not applicable, species not detected in subjects with BMI ≥ 30.

**Table SI7** Bacterial species with differential abundance in stool metagenomes of subjects with Body Fat (BF) ≥ 40% compared to those with BF < 40% at baseline

| Species | Abundance Subjects with BF < 40%at Baseline | | Abundance Subjects with BF ≥ 40% at Baseline | | Fold Change Log2(BF ≥ 40% / BF < 40%) | p-value (Welch's t-test) | p-value (Welch's t-test with FDR correction) |
|---|---|---|---|---|---|---|---|
| | Mean | SD | Mean | SD | | | |
| UBA11524.sp000437595 | 0.7073 | 0.628 | 0.284 | 0.494 | -1.32 | 3.43E-08 | 2.57E-05 |
| Clostridium.sp000435835 | 0.1369 | 0.198 | 0.034 | 0.089 | -2.01 | 5.52E-07 | 0.0002 |
| Dysosmobacter.welbionis | 0.1478 | 0.177 | 0.302 | 0.286 | 1.03 | 2.94E-06 | 0.0007 |
| Schaedlerella.glycyrrhizinilytica_A | 0.0154 | 0.058 | 0.073 | 0.119 | 2.25 | 9.23E-06 | 0.0017 |
| Ventrisoma.faecale | 0.0029 | 0.023 | 0.036 | 0.081 | 3.64 | 7.98E-05 | 0.0120 |
| Mediterraneibacter.torques | 0.3285 | 0.462 | 0.615 | 0.625 | 0.90 | 0.0001 | 0.0138 |
| Choladousia.sp902363665 | 0.2369 | 0.241 | 0.125 | 0.197 | -0.92 | 0.0001 | 0.0138 |
| Fimenecus.sp000432435 | 0.0784 | 0.259 | 0.301 | 0.538 | 1.94 | 0.0001 | 0.0138 |
| Blautia_A.hydrogenotrophica | 0.0313 | 0.089 | 0.091 | 0.138 | 1.54 | 0.0002 | 0.0146 |
| CAG.313.sp003539625 | 0.1873 | 0.439 | 0.030 | 0.142 | -2.63 | 0.0003 | 0.0193 |
| Fimadaptatus.sp900553645 | 0.0582 | 0.108 | 0.016 | 0.064 | -1.86 | 0.0003 | 0.0213 |
| Lachnospira.sp003451515 | 0.1718 | 0.284 | 0.059 | 0.185 | -1.55 | 0.0004 | 0.0233 |
| Flavonifractor.plautii | 0.0741 | 0.155 | 0.169 | 0.233 | 1.19 | 0.0004 | 0.0251 |
| Blautia_A. sp900066205 | 0.1838 | 0.314 | 0.070 | 0.161 | -1.39 | 0.0005 | 0.0288 |
| CAG.170.sp900545925 | 0.1717 | 0.255 | 0.074 | 0.177 | -1.21 | 0.0008 | 0.0399 |
| Clostridium.sp900540255 | 0.2305 | 0.351 | 0.103 | 0.209 | -1.16 | 0.0009 | 0.0399 |
| Enterocloster.sp001517625 | 0.0248 | 0.092 | 0.107 | 0.236 | 2.11 | 0.0009 | 0.0416 |
| Blautia_A.wexlerae | 1.4618 | 0.795 | 1.889 | 1.105 | 0.37 | 0.0011 | 0.0431 |
| Ventrimonas.sp900538475 | 0.0107 | 0.058 | 0.068 | 0.169 | 2.67 | 0.0011 | 0.0431 |
| CAG.302.sp000431795 | 0.1371 | 0.230 | 0.048 | 0.181 | -1.50 | 0.0012 | 0.0466 |
| Blautia_A.sp900066505 | 0.1351 | 0.156 | 0.073 | 0.134 | -0.89 | 0.0013 | 0.0468 |

**Table SI8** Summary of gastrointestinal tolerability questionnaires

| | SBD111 (N = 142) | | Placebo (N = 139) | | |
|---|---|---|---|---|---|
| **Number of subjects reporting GI symptoms** | **Yes** | **No** | **Yes** | **No** | **Pearson's $Chi^2$ p-value** |
| Any symptoms | 43 (30.3%) | 99 (69.7%) | 45 (32.4%) | 94 (67.6%) | 0.71 |
| Mild symptoms | 40 (28.2%) | 102 (71.8%) | 42 (30.2%) | 97 (69.8%) | 0.71 |
| Moderate symptoms | 26 (18.3%) | 116 (81.7%) | 30 (21.6%) | 109 (78.4%) | 0.49 |
| Severe symptoms | 4 (2.8%) | 138 (97.2%) | 13 (9.4%) | 126 (90.7%) | **0.02** |
| **Number of GI symptoms reported per participant** | **Mean** | **SD** | **Mean** | **SD** | **Pearson's $Chi^2$ p-value** |
| Any symptoms | 2.09 | 4.49 | 2.73 | 5.70 | 0.42 |
| Mild symptoms | 1.49 | 3.10 | 1.81 | 3.98 | 0.55 |
| Moderate symptoms | 0.56 | 1.73 | 0.73 | 2.00 | 0.48 |
| Severe symptoms | 0.04 | 0.29 | 0.19 | 0.77 | **0.02** |

**Table SI9** Summary of menopause symptom questionnaires

| | **Baseline** | | | **3 Months** | | | **6 Months** | | | **9 Months** | | | **12 Months** | | |
|---|---|---|---|---|---|---|---|---|---|---|---|---|---|---|---|
| | **SBD111 (N=143)** | **Placebo (N=142)** | **p-value** | **SBD111 (N=135)** | **Placebo (N=126)** | **p-value** | **SBD111 (N=122)** | **Placebo (N=114)** | **p-value** | **SBD111 (N=117)** | **Placebo (N=106)** | **p-value** | **SBD111 (N=114)** | **Placebo (N=107)** | **p-value** |
| Overall MRS score | 9.77 (5.45) | 10.85 (5.66) | 0.1 | 6.18 (4.30) | 7.01 (4.90) | 0.79 | 6.84 (4.22) | 7.32 (4.79) | 0.41 | 6.41 (4.49) | 7.01 (4.19) | 0.53 | 6.71 (4.59) | 7.06 (4.69) | 0.22 |
| Somatic MRS Score | 4.03 (2.20) | 4.30 (2.32) | 0.33 | 2.91 (1.95) | 2.81 (1.85) | 0.14 | 3.12 (2.05) | 3.11 (1.90) | 0.37 | 2.80 (1.94) | 2.97 (1.75) | 0.79 | 2.94 (2.05) | 2.94 (1.93) | 0.34 |
| Psychological MRS Score | 3.15 (2.30) | 3.70 (2.46) | 0.053 | 1.99 (1.95) | 2.66 (2.52) | 0.66 | 2.16 (1.96) | 2.57 (2.27) | 0.65 | 2.03 (1.91) | 2.48 (2.26) | 0.87 | 2.18 (2.01) | 2.48 (2.10) | 0.33 |
| Urogenital MRS Score | 2.59 (2.29) | 2.85 (2.22) | 0.32 | 1.28 (1.57) | 1.54 (1.78) | 0.76 | 1.56 (1.79) | 1.65 (1.78) | 0.58 | 1.58 (1.78) | 1.56 (1.60) | 0.21 | 1.59 (1.82) | 1.64 (1.85) | 0.34 |

Values for SBD111 and Placebo groups represent mean (standard deviation). Overall MRS score is obtained by summing the scores of all 11 items on the MRS scale. Somatic MRS score is obtained by summing the items 'hot flushes and sweating', 'heart discomfort', 'sleep problems', and 'joint and muscular discomfort'. Psychological MRS score is obtained by summing the items 'depressive mood', 'irritability', 'anxiety', and 'physical and mental exhaustion'. Urogenital MRS score is obtained by summing the items 'sexual problems and irritability', 'bladder problems', and 'vaginal dryness'. For baseline, p-values are derived from a two-sided student's t-test. For 3, 6, 9, and 12 months, p-values were obtained via linear mixed effects models.
